# Depression, Anxiety, Somatic symptom and their determinants among High School and Preparatory School Adolescents in Gondar Town, Northwest Ethiopia, 2022.Non-recursive Structural Equation Modeling

**DOI:** 10.1101/2023.01.27.23285096

**Authors:** Zenebe Abebe Gebreegziabher, Rediet Eristu, Ayenew Molla

**Author notes:** Corresponding author (ZA).

## Abstract

**Introduction:** In Developing Countries including Ethiopia, mental health is not only a public concern; but also a developmental issue. Depression and anxiety are the commonest mental health disorders and often somatic symptoms co-exist with them. Adolescents with common mental health problems are associated with increased risk of suicide, future unemployment, and poor quality of life. Little is known about the mental health of adolescents in the Ethiopia. Thus, this study aimed to assess the magnitude and determinants of depression, anxiety, and somatic symptoms among high school and preparatory school adolescents in Gondar town.

**Methods:** Institution based cross-sectional study was conducted from June 8 to 24, 2022. Two-stage stratified random sampling was used to select 1407 high school and preparatory school adolescents in Gondar town. Data were collected through structured and standard self-administered questionnaires. STATA version 16 and AMOS version 21 were used for analysis. Non-recursive structural equation modeling was employed to assess the direct, indirect, and total effects of the predictors. Degree of relationship was interpreted based on adjusted regression coefficients with corresponding 95% confidence interval.

**Results:** Magnitude of anxiety, depression, and somatic symptoms were 25.05% (95%CI: 22.8, 27.5), 28.21 (95% CI: 25.8, 31%), and 25.24(95% CI: 23, 27.6%), respectively. High self-rated academic ability (β=-0.03, 95% CI: -0.065, -0.006) and high perceived social support (β=-0.22, 95% CI: 0-, -0.139) had significant negative effect on anxiety. High levels of depression had a direct positive effect (β= 0.74, 95% CI: 0.508, 1.010) on anxiety. High level of stress had significant direct (β=0.54, 95% CI: 0.293, 0.745) and indirect (β=0.57, 95% CI: 0.379, 0.814) positive effect on anxiety. High level of anxiety was significantly related with high level of depression (β=0.74, 95% CI= 0.483, 1.081). High perceived social support (β= -0.13, 95% CI: -0.229,-0.029), and having a history of death of beloved one within the past six months (β= 0.03, 95% CI: 0.014, 0.256) had a significant direct positive effect on depression. Having medically confirmed chronic illness (β=0.21, 95% CI=:0.114, 0.311), being female (β= 0.06, 95% CI= 0.003, 0.109) and high level of stress (β= -0.06, 95% CI: -0,454, 0.247) had significant indirect effect on depression. Stress (β= 0.86, 95% CI: 0.700, 1.025), anxiety (β=0.66, 95% CI: 0.270, 3.825) and depression (β= 0.96, 95% CI: 0.167, 3.629) were significant predictors of somatic symptoms.

**Conclusions and recommendations:** Magnitude of anxiety, depression, and somatic symptoms were moderate. Self-rated academic ability, physical trauma, school type, sex, stress, ever use of alcohol, perceived social support, death of beloved one, and having medically confirmed chronic illness were independent predictors of anxiety, depression and somatic symptoms. The bidirectional relationship between anxiety and depression was significant. Emphasis should be given to the prevention and management of mental health in the adolescent, particularly targeting adolescents with the aforementioned factors.

## Introduction

Mental health disorder is defined as “syndrome characterized by clinically significant disturbance in an individual’s thought, emotion regulation, or behavior that reflects a dysfunction in the psychological, biological, or development processes underlying mental functioning”[1].

Mental health disorder accounts for 16% of global burden of disease and injuries and more than 80% of people with mental health disorders live in low and middle income countries, with increase in incidence and mortality [2]. Adolescent are especially vulnerable to developing mental illness[3]. In this population mental health disorders is increased by 32.18% from 1999 to 2019 [4]. World health organization(WHO) estimates that 10-20% of adolescents are affected by mental health problem in the world [5]. In low- and middle-income countries, adolescents mental health is not only a major public health challenge, but also a development concern [6]. Human suffering and financial costs associated with mental health disorders are substantial and growing globally [7]. Each year more than 12 billion working days are lost due to mental illness. Between 2011-2030, it is estimated that mental illness will cost the global economy 16 trillion united states dollar in lost economic output more than diabetes, cancer, and respiratory diseases [6]. Depression and anxiety are among the common mental health disorders [8],and often somatic symptom co-exist with them [9, 10].

Depression, which is the second common mental health disorder in adolescents is characterized by loss of interest or pleasure, feelings of guilt or low self-worth, feelings of tiredness, disturbed sleep or appetite, and poor concentration and sadness [11]. In combination with anxiety, it contributes for 45% of the overall burden of disease [12]. In 2018, over one million adolescents died from preventable causes, with depression playing a significant role [13]. In Asia, its magnitude ranges from 24.3% to 57.7% [14] and in Sub-Saharan Africa, it ranges from 15.5% [15] to 45.90% [16, 17]. In Ethiopia, 28% [18] of adolescents in Jimma and 36.2% [19] of adolescents in Aksum were depressed.

Anxiety is characterized by emotional feelings of excessive fear, nervousness, avoiding treats in the environment perceived by them, and physical symptoms such as fast respiration, increased blood pressure, and tightness of the chest [20, 21]. Globally, 301.39 million peoples are affected by anxiety [22]. It is the leading cause of mental disorder in the world, which accounts for about 28.68 million disability-adjusted life years in the global burden of disease (GBD) [23]. Anxiety is the commonest mental health disorder in adolescents [24], with high magnitude in developed as well as in developing countries[25]. In adolescents, it ranges from 42.1% [26] to 80.85% in Asia [27], and in sub-Saharan Africa 29.8% of adolescents had anxiety [3]. A study conducted in Kenya showed that 37.99% of high school adolescents had anxiety [17]. A systematic review conducted among children and youth in Ethiopia revealed that general anxiety disorder ranges from 0.5-23% [28].

Somatic symptom is characterized by a comprehensive list of symptoms such as pain, breathlessness, numbness, palpitation, tiredness, headache, dizziness, and gastroenterological problems. Somatic symptom disorder is a disorder in which individuals excessively or disproportionally think or feel about the symptoms, which result in significant disturbance in daily life [29]. Often depression and anxiety co-exists with somatic symptom [30–32], which is one of the remarkably widespread issues in primary health care and subspecialty settings [10]. Somatic symptoms results in more than half of the patient visit in primary health care [33]. Its magnitude is estimated to be 5 to 7% [34] in general population and 5 to 30 % [35] [36] in adolescent.

Studies showed that age [17], female sex [18, 37–39], being in public school [40], high family academic pressure [41] [42], poor self-rated academic ability [43, 44], alcohol use [45] [46], cigarette smoking [47, 48], stress, [49, 50] [51], somatic symptom[18, 37, 38, 49, 52, 53], and anxiety [54, 55] had a positive relation with depression. While other factors like social support [43, 44, 52, 56] and family education had negative association [53].

Literatures reported that female sex [27, 57], age [17, 26], smoking [48, 58], alcohol use, self-rated academic ability [26, 59], family academic pressure [41]. Stress [60, 61], somatic symptom disorder [62] and depression [63] had positive relationship with anxiety.

Different scholars reported that female sex [64] [65, 66], being in private school [64], physically activity [67, 68], extra school tutoring [69], stress [70] [71, 72], anxiety [65, 73, 74] and depression [65, 75, 76] had significant relation with somatic symptom.

Common mental health disorders in adolescents are associated with severe mental health problems in the future, suicidal ideation, increased risk of drugs and substance abuse, violent behavior, unprotected sex and acquired sexually transmitted infections, school dropouts, future unemployment, and poor quality of life [77–82]. Since it is commonly associated with heart disease, cancer, and diabetes mellitus, depression increases pre-mature death by 40-60% [83]. Somatic symptoms delay the diagnosis of depression and anxiety, because most people seek medical attention for bodily symptoms. Individuals with somatic symptoms experience harm from unnecessary testing and treatment [84].

Despite the fact that mental health is a key part of the sustainable development goals (SDG) which affect every other goals [6], the magnitude of mental health disorders is still high [27]. To decrease the burden of mental illness, targeting adolescents is important; since more than half of mental health problems starts at this age [6], and 50% of adolescent mental health problems will proceed to adulthood [85]. Adolescents in low and middle-income countries are disproportionally affected by mental health disorders; however, there are limited data on the prevalence and determinants of mental illness in Sub-Saharan Africa [86] [87].

In Ethiopia, though mental health services were included in the national health policy, there is a paucity of evidence on the magnitude and determinants of anxiety, somatic symptoms, and their relation with depression. Even studies conducted outside of Ethiopia had methodological flaws, as they used logistic or linear regression to assess multiple mental health outcomes, which cannot address the bidirectional relationship between outcome variables and the indirect effects of predictors. In such cases, non-recursive structural equation modeling, which is a multivariate statistical framework used to measure the complex relationships between different observed and latent variables simultaneously is preferable.

Therefore, this study intended to fill this gap by assessing the magnitude, relationship, and determinants of depression, anxiety, and somatic symptoms among high school and preparatory school adolescents in Gondar town by using non-recursive structural equation modeling. The findings from this study will help the psychologist, psychiatrists, policy makers, students, community and the government to tackle the increasing burden of mental health problems. Moreover, it will be used as an input toward sustainable development and as a baseline data for further research.

## Methods and Materials

### Study design, setting, and period

Institution based cross-sectional study was conducted from June 8 -June 24, 2022 in Gondar town, Amhara Region state, northwest, Ethiopia. It is located 728 kilometers from Ethiopia’s capital city of Addis Ababa, and 180 kilometers from Bahir Dar, the capital city of the Amhara regional state. There are 395, 000 residents [88], 48 healthcare facilities (one comprehensive specialized hospital, eight health centers, one private general hospital, fifteen specialist clinics, fifteen medium clinics, and eight primary clinics). According to the town administrative educational department report, the town has seventeen high and preparatory schools (twelve public and five private), with a total number of 24,308 students. Among them, 2408 students were from private schools (1457 girls and 951 boys) and 21900 students were from public schools (12052 girls and 9848 boys).

### Population

All high and preparatory school adolescents in Gondar town were the source population and all high and preparatory school adolescents in randomly selected schools in Gondar town were the study population.

### Eligibility criteria

All high and preparatory school adolescents in Gondar town who were registered for the second semester of 2021-2022 academic years were included in the study. Night preparatory and high school students and students who were transferred in from other areas after January 8, 2022 were excluded.

### Sample size determination

Sample size calculation for structural equation modeling depends on the model complexity. The general rule of thumb approach for sample size calculation in structural equation modeling ranges from 5 to 20 times per free parameters[89]. In this study, 5:1 ratios were used to get the minimum adequate sample size. Based on the hypothesized model *Fig 1*, a total of 134 free parameter were available(18 variance for exogenous variables), (40 regression coefficients which are; the coefficients between exogenous observed variables and latent variable, and coefficients between latent variables), (32 loading which is the loading between latent variables and indicators except those path coefficients fixed to one (5 path coefficients fixed to one in this study), (37 errors variance for indicator’ variables), (3 covariance between disturbances), (4 disturbance of endogenous latent variables) (***Fig 1***).

**Fig 1:**
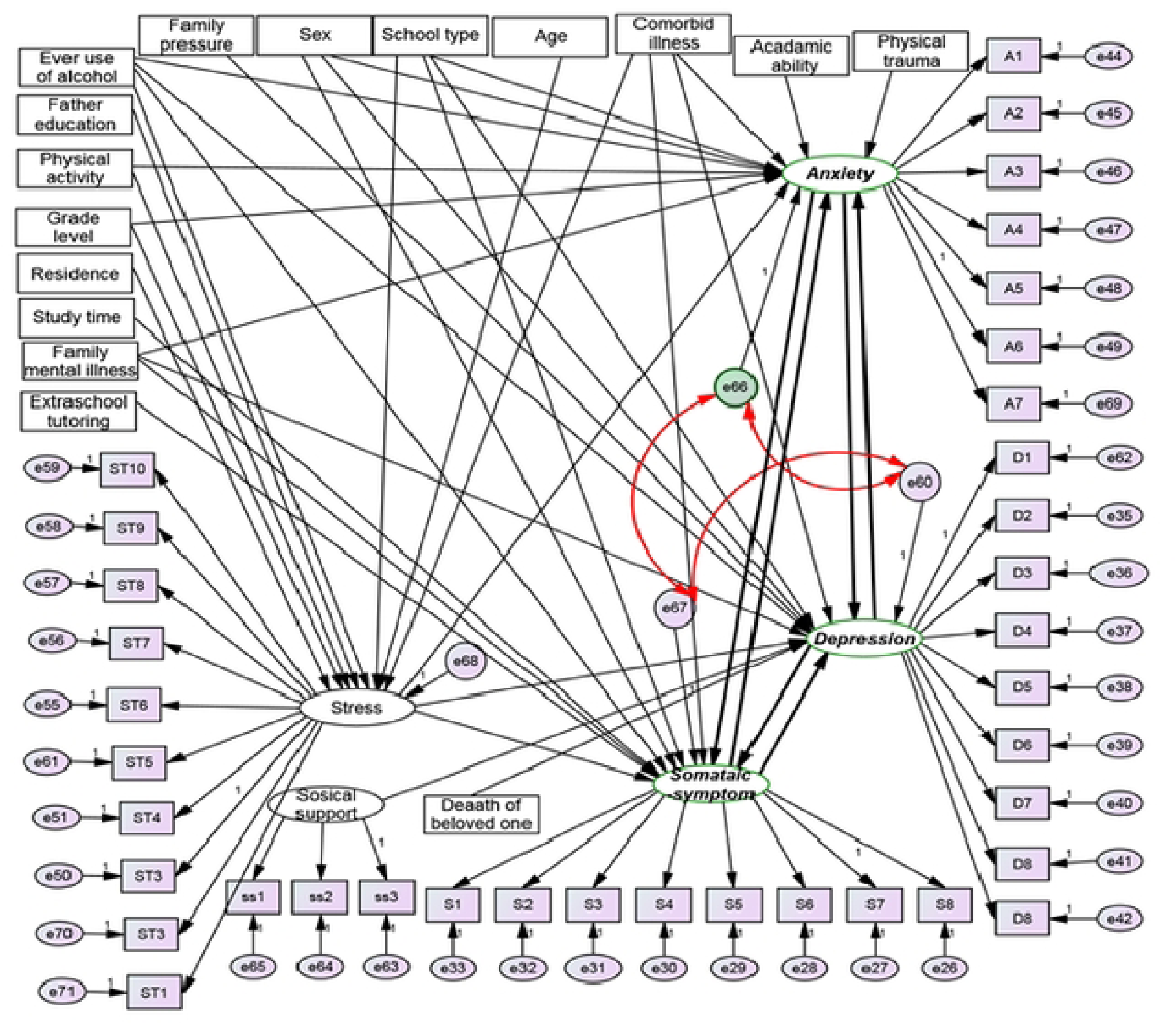
Hypothetical structural equation modeling on the magnitude and determinants of Depression, Anxiety and somatic symptoms among high school and preparatory school adolescents in Gondar town 2022 (both measurement and structural model

Considering 134 free parameters, the sample size was 134*5=670

Since the sampling procedure is multistage sampling with 2 stages, a design effect of 2 was considered.

Therefore, the sample size becomes 670*2 =1340

After adjusting 5% for non-response rate, the final sample size became 1340+1340*5%=**1407**

### Sampling procedure

In this study, two-stage stratified random sampling technique was used to select participants. In Gondar town, there are seventeen high and preparatory schools; of which twelve are public schools (Shinta, Hidar 11, Ayer Tena, Azezo Preparatory, Hidase Teda, Azezo Dimaza, Loza Birhan, Fasiledus Preparatory, Fasiledus General Secondary and Preparatory, Walagi, Anigereb, and Felegi), and five are private schools(Debre selam, Waliya, Kings Acadami, Gondar university community and Abune Samuel). A study conducted in Telangana state, India, showed that adolescents in public schools had a higher proportion of depression than those in private school [40]. As a result, stratification by school type was done and study subjects were proportionally allocated. Then, two schools from private (Walya and Debre selam) and four schools from public (Azezo preparatory school, Fasiledus preparatory school, Shinta, and Anigereb) were selected randomly. After that, heterogeneity by grade level was considered and, again stratification was done by grade level. Students were proportionally allocated in each grade and finally 1407 students were selected randomly by computer generated random sampling (***Fig 2***).

**Fig 2:**
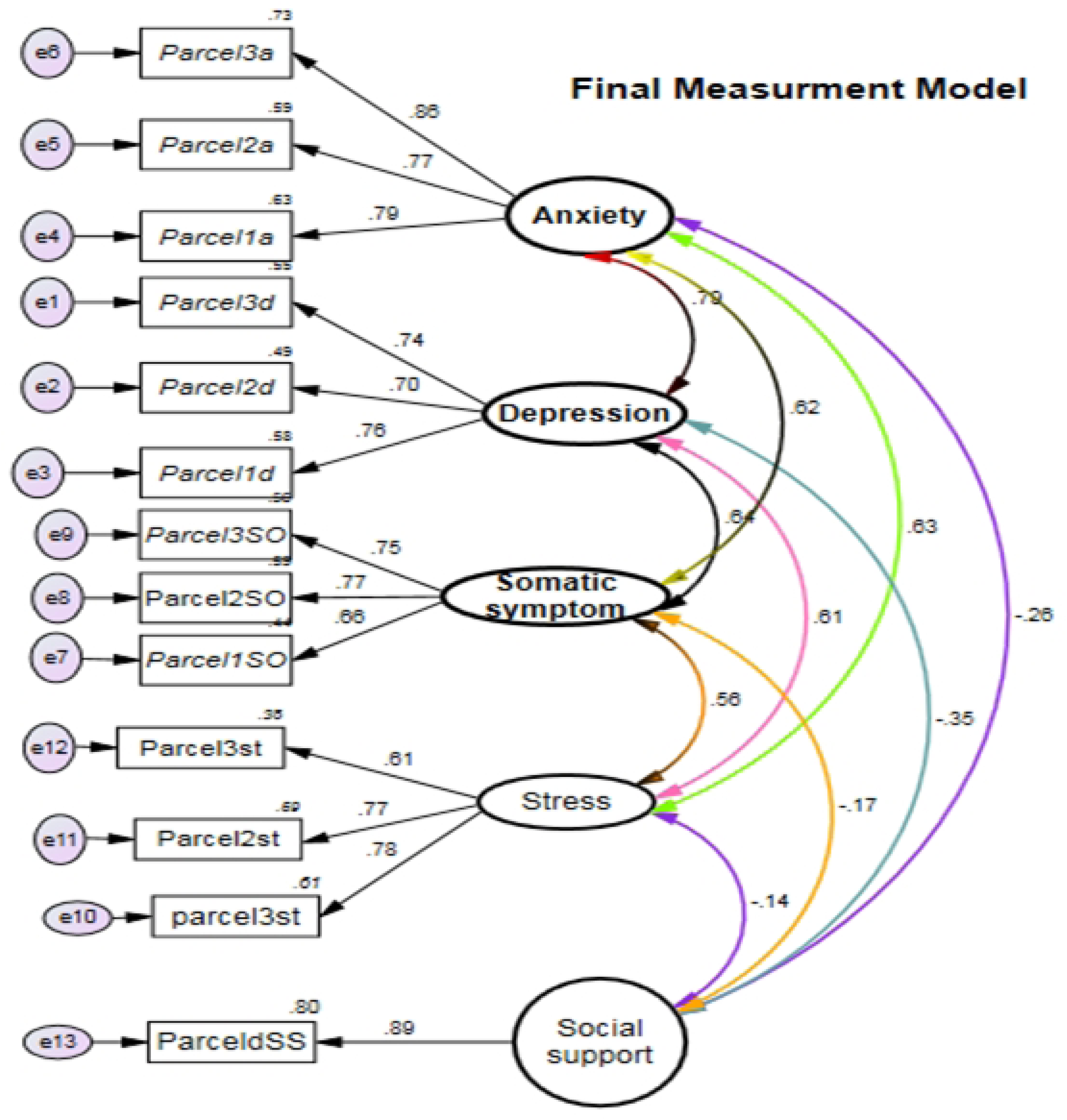
Diagrammatical representation of the sampling procedure that was used to assess the magnitude and determinants of depression, anxiety, and somatic symptoms among high school and preparatory school adolescents in Gondar town, 2022. Where N= total number of students

**Fig 3:**
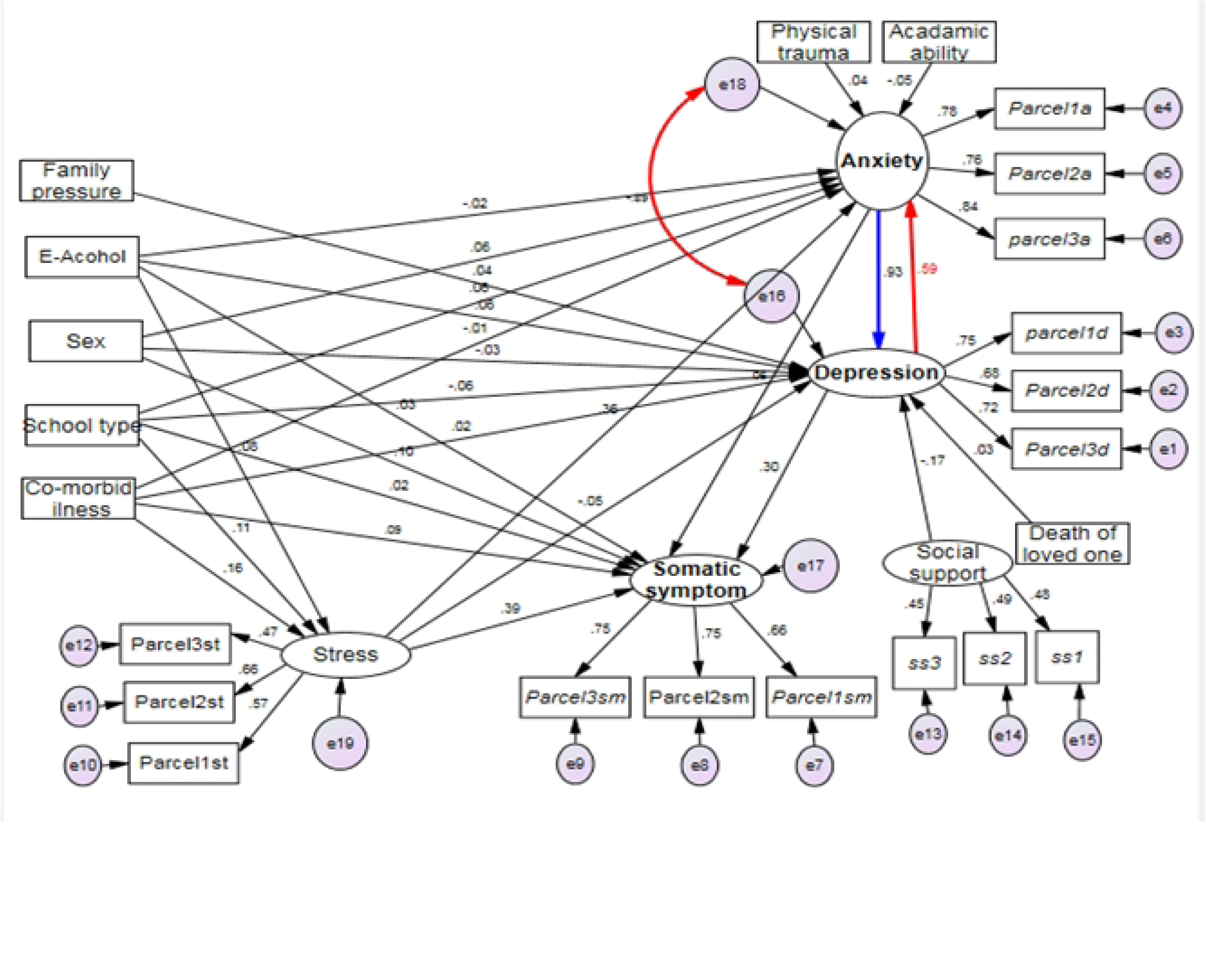
Final measurement model for the constructs of anxiety, depression, somatic symptoms, stress, and social support.

## Variables of the study

### Outcome variables (endogenous constructs)

Depression, anxiety, and somatic symptoms

### Independent variables

#### Observed Exogenous Variables

**Socio-demographic variables**: age, sex, original residence, grade level, school type, father education, and mother education

**Behavior related factors**: physical activity, alcohol use

**Academic related factors**: extra school tutoring, number of study hours per day, self-rated academic ability, and family academic pressure

**Relationships and related factors**: Death of beloved one

**Clinical factors**: family history of mental illness, history of physical trauma, medically confirmed chronic illness

**Exogenous latent variable:** social support

**Endogenous latent variable:** Stress

## Operational definition

**Depression:** Students were classified as having depression, if their total score on the Patients Health Questionnaire (PHQ-9A) was 10 or higher.

Regarding the severity level of depression, students were categorized as those with mild, moderate, moderately severe and severe depression, if they had a total score of 5-9, 10-14, 15-19, and 20-27 respectively [90].

**Anxiety:** Students were classified as anxious if their total score on the general anxiety scale (GAS-7) was 10 or higher [91]. Regarding the severity level of anxiety, they were categorized as having mild 5–9 score, moderate 10–14 scores, and severe anxiety with a score of 15–21 [92].

**Somatic symptom:** Participants were classified as having somatic symptoms when they scored ≥9 in the somatic symptom Scale (SSS-8), which has an optimum sensitivity of 72% and specificity of 59% [93].

Students were classified as having 4 to 7 low, 8 to 11 medium, 12 to 15 high, or 16 to 32 very high somatic symptom severity levels [94].

**Perceived Stress:** Students who have a total perceived stress(PSS-10) scores of 0-13, 14-26, and 27-40 were categorized as having low, moderate, and high perceived stress level respectively [95].

**Perceived social support:** Based on Oslo social support scale(OSS-3), students were categorized as having poor social support 3–8 score, moderate social support 9–11 score, and strong social support with 12–14 score [96].

**Physical activity:** a student who answered 2 days and above for the question in the past one week, how many days did you exercise for at least 60 minutes until we felt sweaty or shortness of breath were categorized as physically active; and those who answered one day and no was categorized as physically inactive [97].

**Ever use of alcohol**: Students who had a history of alcohol drinks (like beer, Tella, Katikala, Wine) at least once in his/her life time were considered as positive for ever use of alcohol [98].

**Current alcohol use**: Individuals who have a history of alcohol drunk in the past 3 months were considered as positive for current alcohol user [98].

## Data collection procedures and tools

Primary data were gathered using structured questionnaire through self-administered questionnaire for those who could see and by face-to-face interviews for individuals who could not see. Four trained data collectors who have a first degree in public health officer were assigned for data collection.

The tool for this study contains 9 domains which are: socio-demographic characteristics, behavioral factors, academic related factors, and relationship related factors, clinical factors, depression, anxiety, somatic symptom, and stress domains.

Depression was assessed using PHQ-9A, it is a depression screening tool with 9-items, each item were measured by Likert scale ranging from 0 “not at all” to 3 “nearly every day” and its total score range from 0 to 27 [99].

Anxiety was assessed using GAS-7, which is a brief measure of anxiety that contains seven items which are rated in four point Likert scales that range from 0 “not at all” to 3 nearly every day”. Its summed score ranges from 0-21 [91].

Somatic symptom was assessed using SSS-8, which had comparable reliability and validity with the patient’s health questionnaire (PHQ-15) [100]. Each item was measured by Likert scale ranging from 0 “not at all” to 4”very much’, a total scoring range from minimal 0 to maximum 32 [94].

Stress was assessed by using Perceived stress scale (PSS-10), it contains 10 items. Each item scored on an ordinal scale of 0–4, where 0 represents “never” and 3 represents “very often”. Individual scores range from 0 to 40 [95].

Perceived social support: Oslo Social Support scale (OSSS-3) was to assess the perceived social support of students. It is 3 items with a Likert scale that ranges from 1 to 4 for the first item (number of people you can count if you face a great personal problem) and 1 to 5 ranges for the rest items, its summed score range from 3 to 14 [96] .

## Data quality and management

To maintain the quality of the data, one day training was given for data collectors on the objective of the study, the technique of data collection, the content of the questionnaire, and the issue of confidentiality. Before the actual data collection, the questionnaire that developed in English language was translated in to Amharic and re-translated in to English by another person to check consistency. Four public health experts and two psychiatrists checked the face validity of the tool. Construct validity and reliability were checked, since the tool for anxiety, somatic symptoms, and stress was not validated among Ethiopian adolescents, and tools validated in other countries may not be reliable and valid in Ethiopia due to socioeconomic, sociocultural, and linguistic differences.

Moreover, a pilot study was also used to check the way that each questionnaire were understood by different participants and modifications were done accordingly based on the feedback. None of the participants answered yes to smoking; as a result, it was excluded from the final study.

The data collectors immediately checked the collected data for completeness and consistency, and early correctional measures were taken. Repeated trials were taken into account for responders who were unavailable at the time of data collection. Data were cleaned, coded, and entered in to Epi-data software. During data entry, missing, outliers and entry errors were checked manually by going back to the checklist. After data entry, data recording and missing value management were considered accordingly.

### Reliability and validity of the tool

To validate the tool, external participatory pilot study was conducted among 201 students, 177 from Azezo Dimaza General High and Preparatory school (public), and 24 from University of Gondar Community High school (private).

Reliability of the tool was checked by using Cronbach alpha and the tool was reliable, since the Cronbach alpha values were above 0.7 for both overall and particular factors except for social support (***Table 1***). For social support, since it had small number items (<5), average inter-item covariance (AIIC) value was used, thus was 0.4, which in the acceptable range (0.2 to 0.4) [101].

The convergent and discriminate validity of the tool was checked. Even if the Average Variance Extracted (AVE) was below 0.5, the convergent validity was maintained. Since, all standardized factor loadings were significant (p value <0.05) and composite reliability was above 0.6 [102] (*Table 1*). The discriminate validity of the tool was also evidenced, since all inter-factor correlations were less than the square root of AVE for each factor (***Table 1***).

**Table 1:**
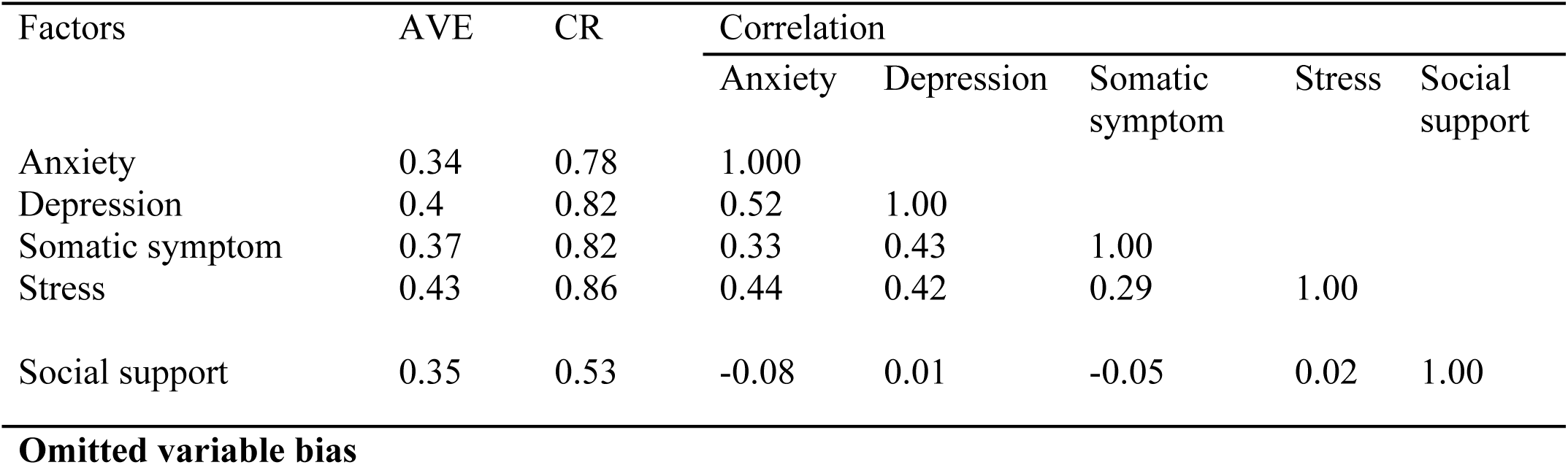
Average variance extracted, composite reliability, and Spearman rank correlation of tools used to measure anxiety, depression, somatic symptoms, stress, and social support among high and preparatory school adolescents in Gondar town northwest Ethiopia 2022.

| Factors | AVE | CR | Correlation |  |  |  |  |
| --- | --- | --- | --- | --- | --- | --- | --- |
|  |  |  | Anxiety | Depression | Somatic symptom | Stress | Social support |
| Anxiety | 0.34 | 0.78 | 1.000 |  |  |  |  |
| Depression | 0.4 | 0.82 | 0.52 | 1.00 |  |  |  |
| Somatic symptom | 0.37 | 0.82 | 0.33 | 0.43 | 1.00 |  |  |
| Stress | 0.43 | 0.86 | 0.44 | 0.42 | 0.29 | 1.00 |  |
| Social support | 0.35 | 0.53 | -0.08 | 0.01 | -0.05 | 0.02 | 1.00 |

Omitted variable bias (OVB) was also checked in the pilot study. It occurs when a statistical model eliminates one or more independent variables that are correlated with one or more of the included independent variables and are determinants of the dependent variable. Ramsey regression equation specification error test (RESET) was used to check OVB. Ramsey (RESET) values above 0.05 indicates the absence of omitted variable bias [103]. In our case, the minimum Ramsey RESET p value was 0.2 (*Table **2***). Hence, there was no significant omitted variable bias.

**Table 2:** Ramsey RESET for anxiety, depression, and somatic symptoms considering all predictors

| Dependent variable | Ramsey RESET test<br>P-value |
| --- | --- |
| Anxiety | 0.83 |
| Depression | 0.61 |
| Somatic symptom | 0.20 |

### Data processing, model building and analysis

Collected data were coded and entered in to Epi-data software version 4.6, and exported to STATA version 16 and AMOS version 21 for further analysis. Descriptive analyses were done and text, tables, graphs, charts, and figures were used for data summarization. Non-recursive structural equation modeling was employed to assess the complex relationship between different latent and observed variables.

Structural equation modeling contains two components which are measurement model and structural model [104]. **Measurement model:** Is a confirmatory factor analysis model for grouping multiple indicators in to some constructs [104]. In this study, the measurement model was used to assess the relationship between anxiety, depression, somatic symptoms, stress, and social support with their indicators. Indicators were reflective. i.e. path is directed from the construct to the indicators.

**Structural model**: It was used to assess the interrelationship between latent variables and the relationship between latent variables and observed predictors [104]. In this study, the structural components were the interrelationship between depression, anxiety, and somatic symptoms, and their relation to other latent and observed predictors.

### Assumptions in non-recursive structural equation modeling

**Specified model**: Model specification is the first step in structural equation modeling. It is a statement of the theoretical model in terms of equations or a diagram [104]. In this study, we specified the model after extensive literature searching and reading as presented in ***Fig 1***.

**Identified Model**: There are three levels of identification in the structural equation model: under-identified, just identified, and over-identified. To precede with the given model, the model should be over identified or just identified [104]. For non-recursive models, instrumental variables(IV) are mandatory to estimate the parameters [104]. Instrumental variables are methodological devices used to resolve problems of endogeneity; which occurs when an explanatory variable correlates with the error term of the dependent variable. All non-recursive models suffer from endogeneity problems and need IVs. In an equation system as IV→X→Y, IV is any variable that has a strong and significant direct effect on X, but is not correlated with the disturbance term of Y [104].

In this study, that variables affect anxiety directly, but does not affect other latent variables directly are instrumental for anxiety i.e., perceived academic ability and history of physical trauma. For depression, perceived social support and death of beloved one are an instrumental variables and extra school tutoring and average study time per day are an instrumental variables for somatic symptom disorder. In our model (***Fig 1***), there are three reciprocal loops and all disturbances of latent variables involved in the feedback loops are correlated. Therefore, the identification of non-recursive structural equation modeling with complete correlation of error terms was used. There are three ways of identification in non-recursive structural equation modeling; which are: presence of unique instrumental variables, order condition, and rank condition. Among this, only the rank condition is sufficient for identification, and the rest two are necessary, but not sufficient [104].

#### A unique instrumental variable

In this study, all endogenous variables in the reciprocal loop had unique instrumental variables.

#### Order condition

It is a necessary but not sufficient condition for model identification. The number of excluded variables for that specific endogenous variable minus the total number of endogenous variables minus one should be greater than or equal to 0 [104]. In our model, to satisfy order condition, each endogenous variable in the recursive loop should have at least 3-1 =2 excluded variables. Where, 3 is the number of endogenous variables in non-recursive loop. In this study, all endogenous variables in the non-recursive have more than two excluded variables. For Depression, anxiety, and somatic symptom disorder, 9, 8, and 11variables were excluded. Therefore, model can full fill requirement of order condition.

#### Rank condition

In this particular study, there are 3 endogenous variables in the non-recursive loop and 17 exogenous variables and one endogenous variable out of the feedback loop (***Fig 1***). Rank Conditions were done using the system matrix. To formulate the system matrix, a separate equation for each endogenous variable in the feedback loop is mandatory. Therefore, the equations for depression, anxiety, and somatic symptom disorder are presented as follows

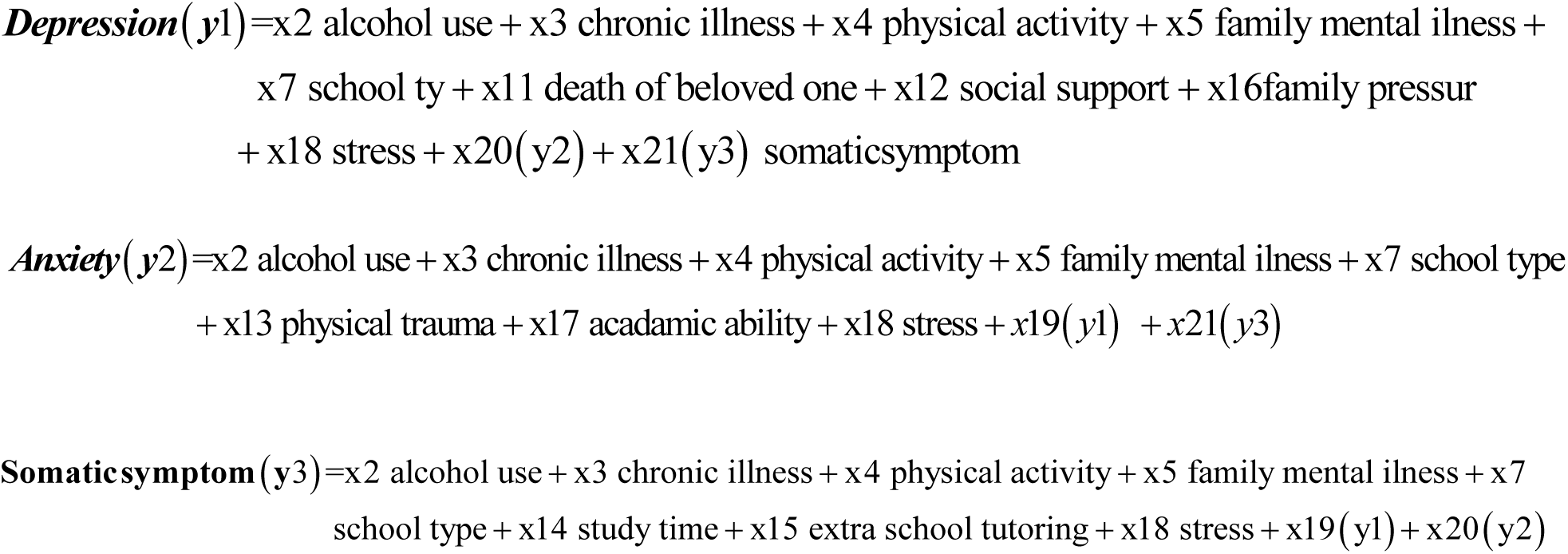

Rank conditions in the system matrix below, begins by constructing a system matrix based on the above equations. Endogenous variables are presented in the row and all variables are presented in the column. In each row, zero or one appears in the column that corresponds to that row. One indicates that the variable represented by the column has a direct effect on endogenous variable in that row or on the endogenous variable itself, 0 indicates the excluded variables for that specific endogenous variable in that row [104].

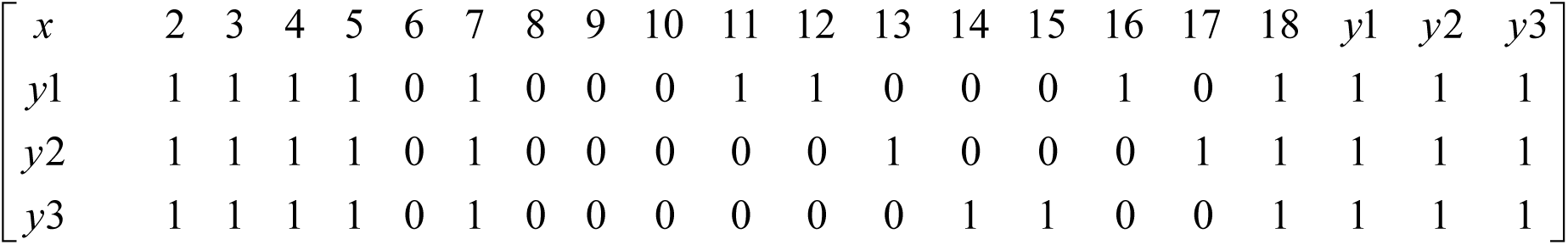

The following steps were followed for identification of rank condition

1. Start with the first row of the system matrix. Cross out all entries in that row and cross out any column in the system matrix with a one in that same row.
2. Simplify the reduced matrix further by deleting any row with all zeros entries; delete any row that is an exact duplicate of another or can be reproduced by adding other rows (i.e., it is a linear combination of other rows). The number of remaining rows is the rank.

If the rank of the reduced matrix is larger than or equal to the sum of the endogenous variables in the recursive loop minus one, then the rank condition is satisfied [104].

The rank condition for this particular study became

For depression (y1), after you cross the entire first row and the entire column which contains 1 in the row:

The reduced matrix becomes:

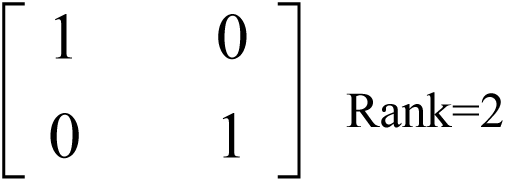

For anxiety (y2), after you cross the entire second row and the entire column which contains 1 in that row:

The reduced matrix becomes

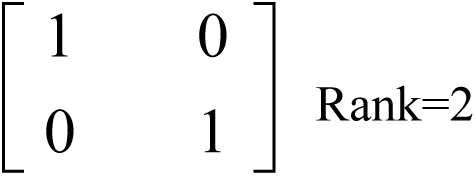

For Somatic symptom (y3), after we cross the entire third row and the entire column which contains one in that row:

Reduced matrix becomes

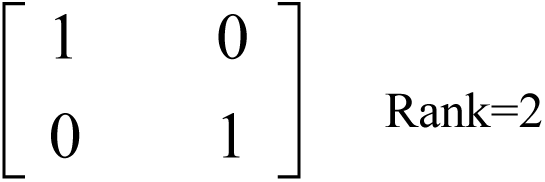

The hypothesized model was identified since all endogenous variables have a rank of two, which is equal to the number of endogenous variables in the feedback loop minus one.

**Multivariate normality, outliers, and missing:** Full information maximum likelihood estimation, which assumes multivariate normality for the joint population distribution of the endogenous variables, given the exogenous variables, was used. Multivariate normality was assessed using Mardia’s kurtosis and its critical ratio. Absolute values of 7 for Mardia’s kurtosis and its critical ratio above 5 were used to declare non-normality [104]. The data were not normally distributed. Although item parceling and outlier deletion were tried to handle it, there was no improvement. As a result, bootstrap maximum likelihood estimation with 3000 samples was used.

Mahalanobis distance(MD), which measure the distance relative centroid, was used to check multivariate outliers [104]. MD p values less than 0.001 based on chi-square distribution were used to declare observation as multivariate outlier [105]. There were 62 observations with Mahalanobis distance p-value less than 0.001. The data were examined to see whether they were actually outliers or caused by data entry errors. However, it was not due to data entry error, and as mentioned above, multivariate normality did not improve significantly with their exclusion. So, those outlier observations were kept in order to maintain the study’s power.

From the total observation, there were six observations with missing values. All missing values were from predictors and had no relationship with the outcome variable; thus, list-wise deletion was performed because they constituted less than 5% of the total sample and were considered missing completely at random [106].

**Sample size adequacy and strength of correlation:** The critical ratios for Bartlett’s test of sphericity in this study were high and it was significant for all constructs. KMO was above 0.7 for the overall, as well as for specific constructs. Hence, the sample is sufficient, the population matrix is not identity, and factor analysis was evidenced[104] (**Table 3**).

**Table 3:** KMO and Bartlett’s test of sphericity to check sampling adequacy and strength of correlation for the measurement of anxiety, depression, a somatic symptoms among high and preparatory school adolescents in Gondar town, 2022.

| Factors | KMO | Bartlett's tests of sphericity |  |
| --- | --- | --- | --- |
|  |  | Chi-square | P value |
| Anxiety | 0.88 | 3100 | 0.0001 |
| Depression | 0.87 | 2391 | 0.0001 |
| Somatic | 0.85 | 2155 | 0.0001 |
| Stress | 0.86 | 3354 | 0.0001 |
| Overall | 0.94 | 13938 | 0.0001 |

**Independency of observations**: The intraclass correlation coefficients (ICC) were used to test the independence of observations. School type and grade level were employed as clustering variables because students are nested in grade level, grade level is nested within schools, and schools are nested in school type. Multilevel SEM was generally taken into consideration at a cut-off value of 0.1 and higher. ICC was less than 0.1 in our study S1 Table1 2, and the usual structural equation model was preferred.

**Common Method Bias:** Common method bias (CMB) occurs when the instrument rather than the actual predispositions of the respondents cause variations in responses. Harman’s one-factor test was used to check common method bias (CMB). The absence of CMB was determined by the overall variance recovered by one factor being below 50% [107]. There was no CMB in this investigation since the overall variance recovered by one factor was 25.2%, which is below the suggested cutoff.

**Strong and valid instrumental variables**: Strength and validity of instrumental variables were checked by using Cragg-Donald Wald F statistic and Sargan Hansen test, respectively. Sargan Hansen test P-value greater than 0.05 was used to ponder valid instruments. F test values above 10 were used as a cut of point to declare strong IVs [108]. In the present study, the strength and validity of the instrumental variables between anxiety and somatic symptoms, and between somatic symptoms and depression were invalid and weak. Deleting a non-recursive path is one treatment for a weak and faulty instrument. As a result, the non-recursive paths from somatic symptoms to anxiety and depression were excluded. Finally, identified model with valid and strong instrumental variables in non-recursive loop was preserved S1 Table1 3.

## Model fitness evaluation and model comparison

**Chi-square:** Chi-square is the inferential goodness of fit index for structural equation modeling. It test is an accept–support test where the null hypothesis represents the researcher’s belief that the model is correct; thus, a failure to reject the null hypothesis, or the absence of statistical significance (p ≥ .05) suggests a good fitted model. Chi-square test is sensitive to sample size; in a large samples it may be below 0.05 in good fitted model [104]. In our study, chi-squares adjusted for degrees of freedom (CMIN/DF) were used, CMIN/DF values 1 to 5 were used to declare acceptable fit [109].

**Rooted mean square error of approximation:** it is the badness of fit index. Values approximate to zero indicate a small discrepancy between the sample implied and the model implied covariance matrix, as its value increase, the model fit becomes poorer. RMSEA departure from close or approximate fit unlike chi-square, measure departure from the exact fit. In this model, a value of 0.05 and below were used to declare good fitted model [110].

**Comparative fit index (CFI):** is one of the relative fix indexes. It was developed as an alternative to the normed fit index (NFI), given that NFI has been shown to be underestimated when small samples are used. 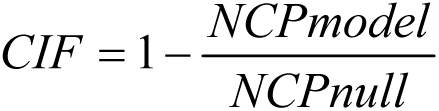, where NCP is non-central parameter, its value ranges from 0 to 1 with values above 0.9 were considered to indicate acceptable fit [110].

**Model comparison**: Model comparison was done using those fit indexes mentioned above and Akaike information criteria (AIC). Finally, a model with minimum information criteria, and RMSEA, and large CFI was selected as a good fitted model

## Ethical consideration

Ethical approval was obtained from the Institutional Review Board (IRB) of University of Gondar Institute of public Health. The ethical approval letter was submitted to Gondar town administrative educational department and to all selected high schools and preparatory schools. Permission was obtained from school directors. Parental informed consent was waived as the research had no/ minimal risk [111]. Following permission from school directors, a detailed participant information sheets were given to each student, and they completed an assent form to indicate a willingness to participate.

To ensure privacy and confidentiality, all data was kept in a locked cabinet at the investigator’s home. Collected data were used for the study purpose only and any personal identifiers were not mentioned. They were told about the benefits and risk objectives and their right to refuse/ discontinue the study at any time without any penalty or loss of benefit.

## Results

### Socio-demographic characteristics and clinical factors of respondents

Out of 1407 randomly selected individuals, 1379 completed the questionnaire; hence, the response rate was 98 %. The mean age of the respondents was 17.23 years with a standard deviation of ±1.25 years. Majority, 1238(89.78%) of the respondents were from public schools and regarding residence two-thirds 915(66.35%) of the respondents were from an urban areas. Around half, 654(47.43%) of the respondents reported moderate stress, while 72 (5.22%) reported high perceived stress. Majority, 1237(89.7%) of the respondents had not a history of chronic illness, and less than one-tenth 101(7.32%) of the respondents had a family history of mental illness **(*Table 4***).

**Table 4:**
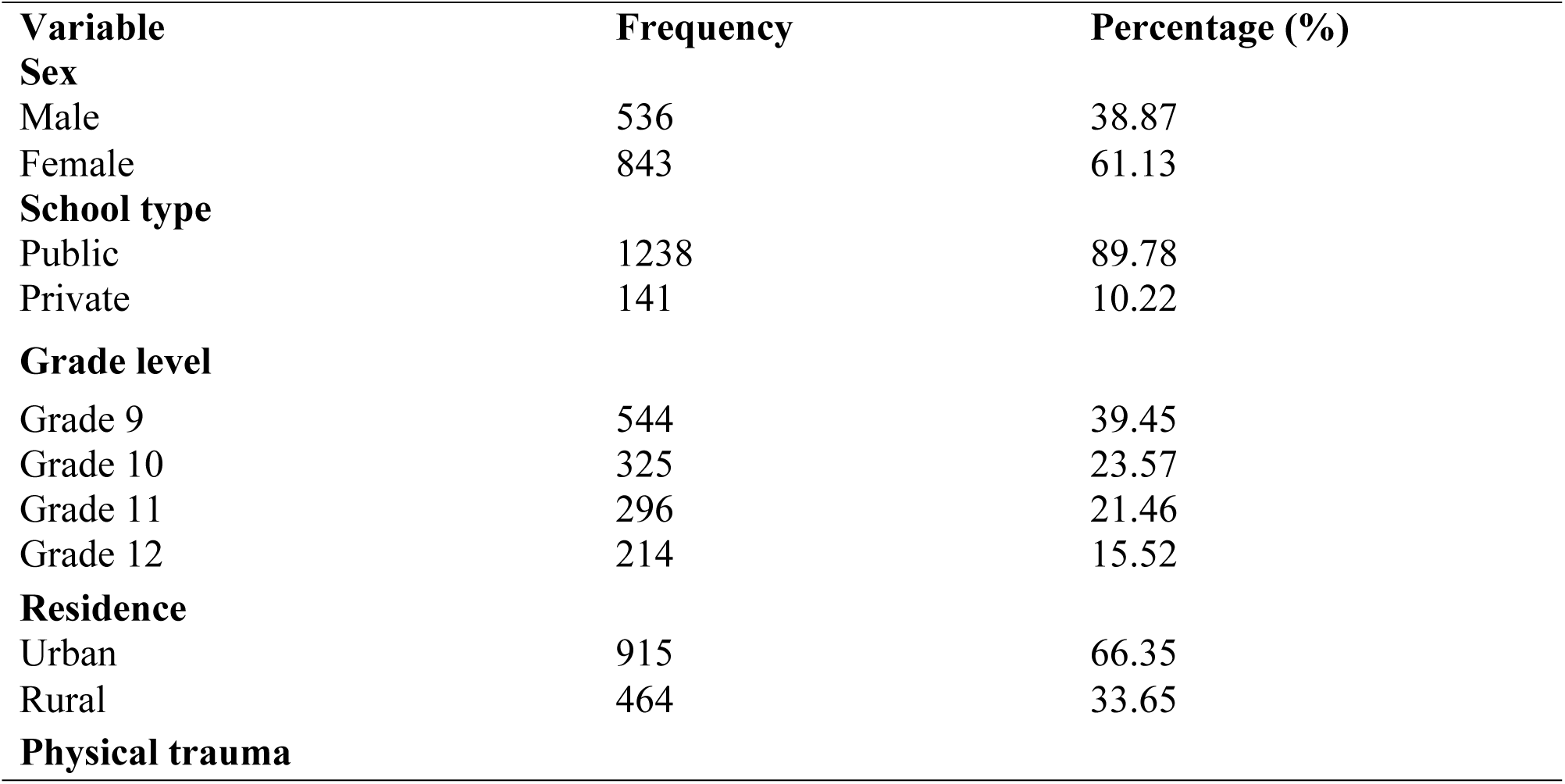
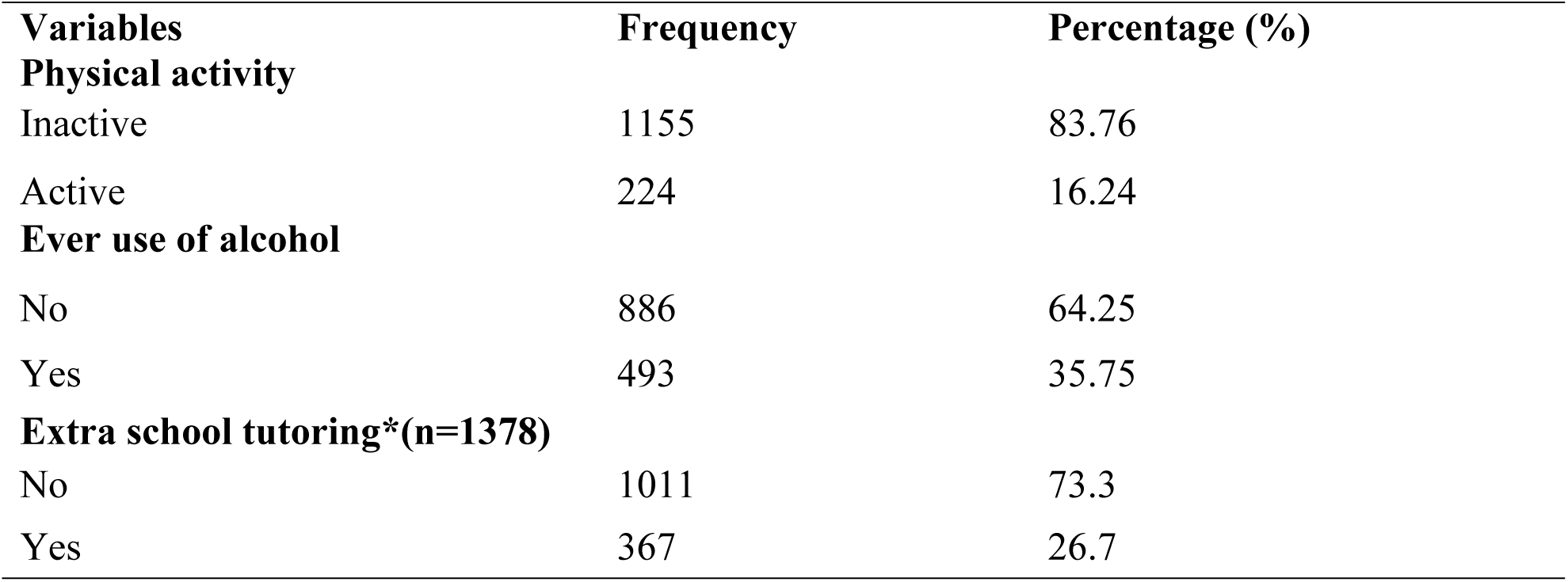
Socio-demographic and clinical characteristics of high and preparatory school adolescents in Gondar town, 2022 (n=1379)

### Behavioral, academic, and relation related factors

More than one-third, 493(35.75%) of the respondents had a history of alcohol drinking. Regarding physical activity, only 224(16.24%) of the respondents were physically active. Majority, 656(47.57% of the respondents had good self-rated academic ability .When we see perceived social support, two-fifths, 564(40.9%) of the respondents had low perceived social support (*Table 5*).

**Table 5:**
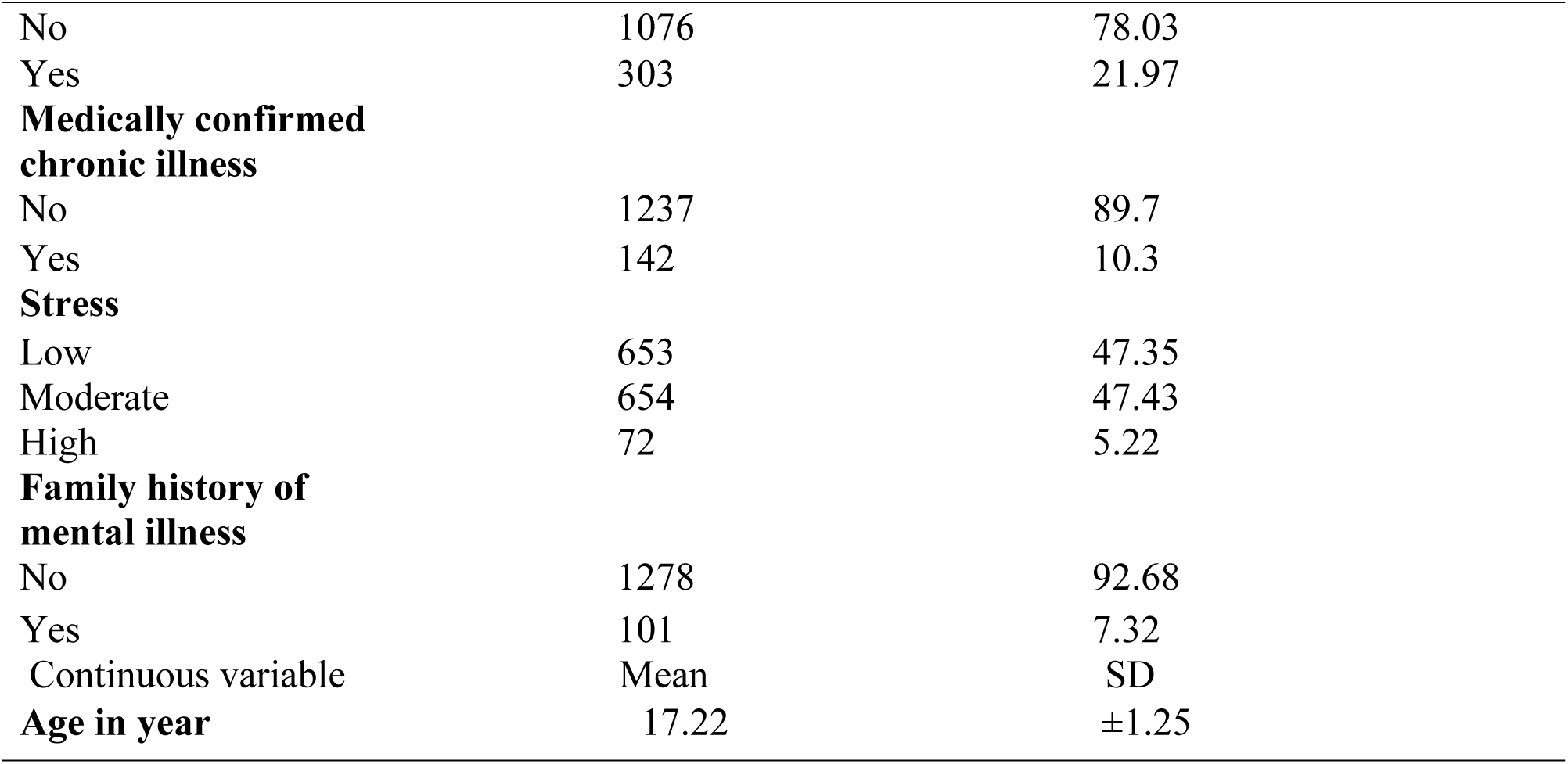
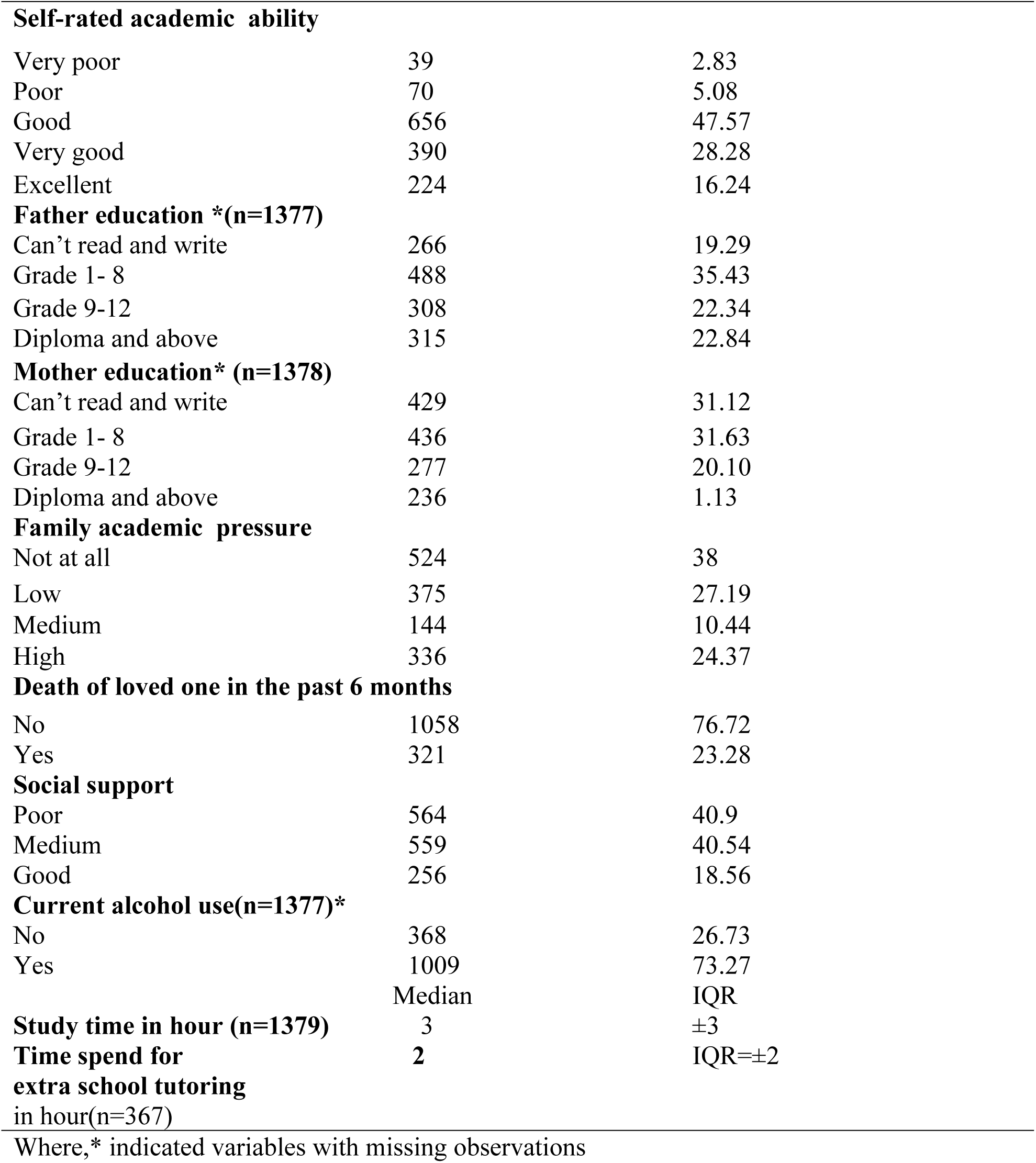
Behavioral, academic, and relation related factors of high preparatory school adolescents in Gondar town, 2022 (n=1379).

### Magnitude of depression

In this study, the overall magnitude of self-reported depression was 28.21(95% CI= 25.8, 31%). Regarding the level of severity, 34.8% had mild, 18.5% had moderate, 7.72% had moderately severe and 2.04% of respondents had severe depression.

More than one-third (34.8%) of participants had a feeling hurting oneself or better off dead. More than two-thirds of the respondents (68.3%) had little interest or pleasure in doing things and more than half of the participants (55.69%) had a feeling of hopelessness S1 Table1 4.

### Magnitude of Anxiety

The overall magnitude of self-reported anxiety in this study was 25.05% (95%CI: 22.8, 27.5%). Regarding the severity level of anxiety, 34.08% of respondents had mild, 16.82% moderate, and 8.27 % had severe anxiety.

Among the total respondents, 16.61% worried too much about different things; nearly every day and 11.54 %could not able to stop worrying too much nearly every day. More than half (58.38%) of the respondents had a feeling of anxious or nervousness S1 Table1 5.

### Magnitude of somatic symptoms

In this study, the overall magnitude of self-reported somatic symptom disorder was 25.24 % (95% CI= 23, 27.6%). Regarding the level of severity among the total respondents, 27.27 % had mild, 15.88% moderate, 8.19% high, and 6.38% very high somatic symptom respectively.

When you see the response to specific indicators, around two-thirds (65.42%) of the participants had headache. More than one half (53.68%) of the respondents had dizziness and 35.97% of the respondent had abdominal pain or gastro intestinal problem S1 Table1 6.

### Confirmatory Factor Analysis

Confirmatory factor analysis (CFA) was computed to test the measurement model. The model fit measures (CMIN/DF, CFI, TLI, RMSEA) were used to assess model overall goodness of fit. In the hypothesized measurement model S1 Fig 1, the goodness of fit was below the acceptable level: CMIN/DF=5.10, TLI=, CFI=0.81 and RMSEA=0.05. To improve the fit of the model, factorial item parceling were done [112] S1 Table1 7. After that, the model fits were improved. In the final measurement model *Fig **1***, all goodness of fit values were in the acceptable level (CMIN/DF=2.60, CFI=1.00, TLI=1.00 and RMSEA=0.00). Construct reliability was assessed by using Cronbach’s alpha and it were above the required level except for social support. For social support, since it had low number item (<5), average inter-item covariance (AIIC) values were used, the value in this study was 0.31, which in the acceptable range (0.20 to 0.40) [101]. Thus, the constructs reliability of the tool was kept up (*Table 6*).

**Table 6:** reliability and model fit for the final measurement model for the constructs of anxiety, depression, somatic symptoms, stress, and social support.

| Conditions | Cronbach alpha | CMIN/DF | CFI | TLI | RMSEA |
| --- | --- | --- | --- | --- | --- |
| Anxiety | 0.84 | 2.6 | 1.0 | 1.0 | 0.0 |
| Depression | 0.79 |  |  |  |  |
| Somatic symptom | 0.77 |  |  |  |  |
| Stress | 0.80 |  |  |  |  |
| Social support | 0.46 |  |  |  |  |
| **Over all** | **0.90** |  |  |  |  |

**Key:** a =anxiety, d= depression, SO=somatic symptoms, st=stress, SS= social support, e =error term, and bidirectional arrow indicates covariance

### Model selection for the hypothesized model containing both measurement and structural model

For the model containing the structural and measurement parts, model comparisons were done using Akaike information criteria (AIC), CMIN/DF, TLI, CFI, and RMSEA. Model with only significant variables with significant disturbance after parcel; which had small AIC, small CMIN/DF, and RMSEA, and large CFI and TLI was selected as a good fitted model (*Table 7*).

**Table 7:** Model selection for a model containing both measurement and structural parts for anxiety, depression, and somatic symptoms among high and preparatory school adolescents in Gondar town, 2022.

| Models | AIC | CMIN/DF | CFI | TLI | RMSEA | Remark |
| --- | --- | --- | --- | --- | --- | --- |
| Model with all predictors and covariance after parcel | 2468 | 6.8 | 0.8 | 0.7 | 0.1 |  |
| Model with only significant variables with significant disturbance after parcel | 951 | 3.00 | 0.94 | 0.92 | 0.027 | selected |

### Factors related with anxiety, depression and somatic symptom

Self-rated academic ability, perceived social support, physical trauma, death of a loved one, ever use of alcohol, having medically confirmed chronic illness, sex, family pressure, school type, and one endogenous variable stress were all significantly related to anxiety, depression, and somatic symptoms, either directly or indirectly. There was a significant bidirectional relationship between anxiety and depression (***Fig 4***, ***Table 8***, ***Table 9***, ***Table 10***).

**Fig 4:**
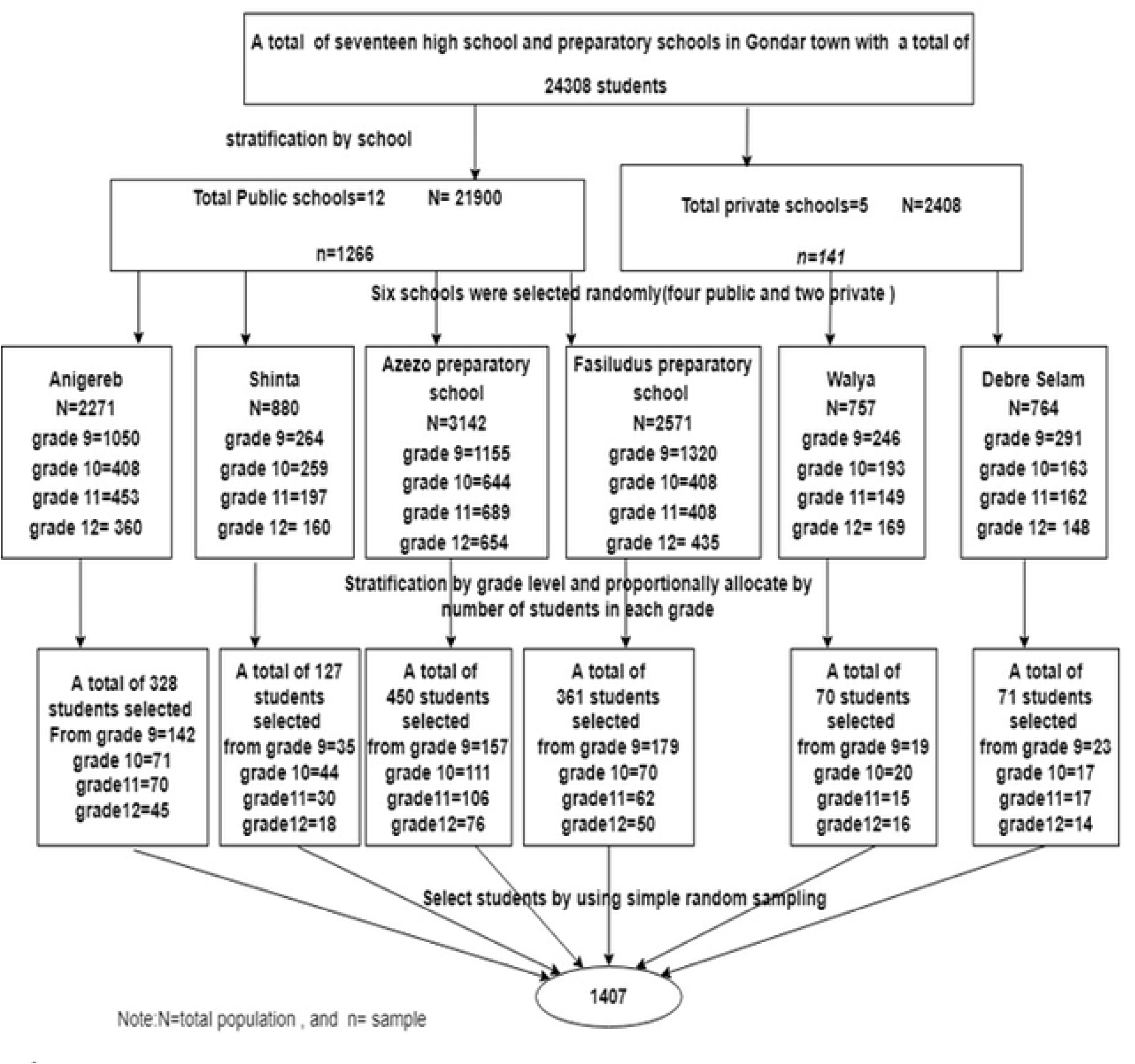
Standardized SEM for factors related with anxiety, depression, and somatic symptom among high and preparatory school adolescents in Gondar town, Northwest, Ethiopia, 2022.

**Table 8:** direct, indirect and total effects of socio-demographic, behavioral, relationship related factors, stress and depression on anxiety among adolescents in Gondar town, Northwest Ethiopia, 2022.

| Variables | Direct effect | Indirect effect | Total effect |
| --- | --- | --- | --- |
| DV: Anxiety | Estimate (95% CI) | Estimate (95%CI) | Estimate (95% CI) |
| Self-rated academic ability | -0.03[-0.065, 0.006 ] * | ---- | -0.03[-0.065, 0.006 ]* |
| Physical trauma | 0.06[0.014, 0.125]* | ---- | 0.06[0.014, 0.125] * |
| Social support | ---- | -0.22[-0.304,-0.139] * | -0.22[-0.304,-0.139] * |
| Death of loved one | ---- | 0.06[0.010, 0.122] * | 0.06[0.010, 0.122] * |
| Ever use of alcohol | -0.03[0.088, 0.035] | 0.14[0.071, 0.213] * | 0.11[0.041, 0.188] * |
| History of chronic illness | -0.02[-0.124, 0.078] | 0.27[0.165, 0.384] * | 0.25[0.132, 0.371] * |
| Sex | 0.08[0.019, 0.135] * | 0.04[0.004, 0.092] * | 0.12[0.059, 0.179] * |
| School type | 0.12[0.017, 0.226] * | 0.16[0.056, 0.269] * | 0.28[0.159, 0.415] * |
| Family academic pressure | ---- | 0.03[0.012, 0.050] * | 0.03[0.001, 0.50] * |
| Stress | 0.54[0.293, 0.745] * | 0.57[0.379, 0.814] * | 1.10[0.955, 1.264] * |
| Depression | 0.74[0.508, 1.010] * | ---- | 0.74[0.508, 1.010] * |

Where \*Indicates CI did not include a null value ( $\beta=0$ ), DV= dependent variable, SE= standard
error, CI=confidence interval

**Table 9:** direct, indirect and total effects of socio-demographic, behavioral, relationship related factors, stress and anxiety on depression among adolescents in Gondar town, Northwest Ethiopia, 2022. **Unstandardized estimate.**

| Variables | Direct effect | Indirect effect | Total effect |
| --- | --- | --- | --- |
| **DV: Depression** | Estimate (95%CI) | Estimate(95% CI) | Estimate (95%CI) |
| Social support | -0.13[-0.229,-0.029 ]* | ---- | -0.13[-0.229,-0.029 ]* |
| Death of loved one | 0.03[0.004, 0.087 ]* | ----- | 0.03[0.004, 0.087 ]* |
| Self-rated academic ability | ---- | -0.05[-0.083, -0.031]* | -0.05[-0.083, -0.031]* |
| Physical trauma | ---- | 0.10[0.037, 0.166 ]* | 0.10[0.037, 0.166 ]* |
| Ever use of alcohol | 0.06[-0.004, 0.112] | 0.08[0.029, 0.139 ]* | 0.14[0.073, 0.201]* |
| History of chronic illness | 0.04[-0.053, 0.124] | 0.17[0.085, 0.265 ]* | 0.21[0.114, 0.311]* |
| Sex | -0.03[-0.099, 0.025] | 0.09[ 0.042,0.162 ]* | 0.06[0.003, 0.109]* |
| School type | -0.09[-0.194, -0.004]* | 0.20[0.113, 0.320] * | 0.11[0.016 , 0.207]* |
| Family pressure | 0.02[-0.004, 0.040 ] | 0.02[0.002, 0.043 ]* | 0.04[0.016, 0.060]* |
| Stress | -0.06[-0.454, 0.247] | 0.82[0.530, 1.250 ]* | 0.76[0.642, 0.900 ]* |
| Anxiety | 0.74[0.483, 1.081*] |  | 0.74[0.483, 1.081*] |

Where \*Indicates CI did not include a null value ( $\beta=0$ ), DV= dependent variable, SE= standard error, CI=confidence interval

**Table 10:**
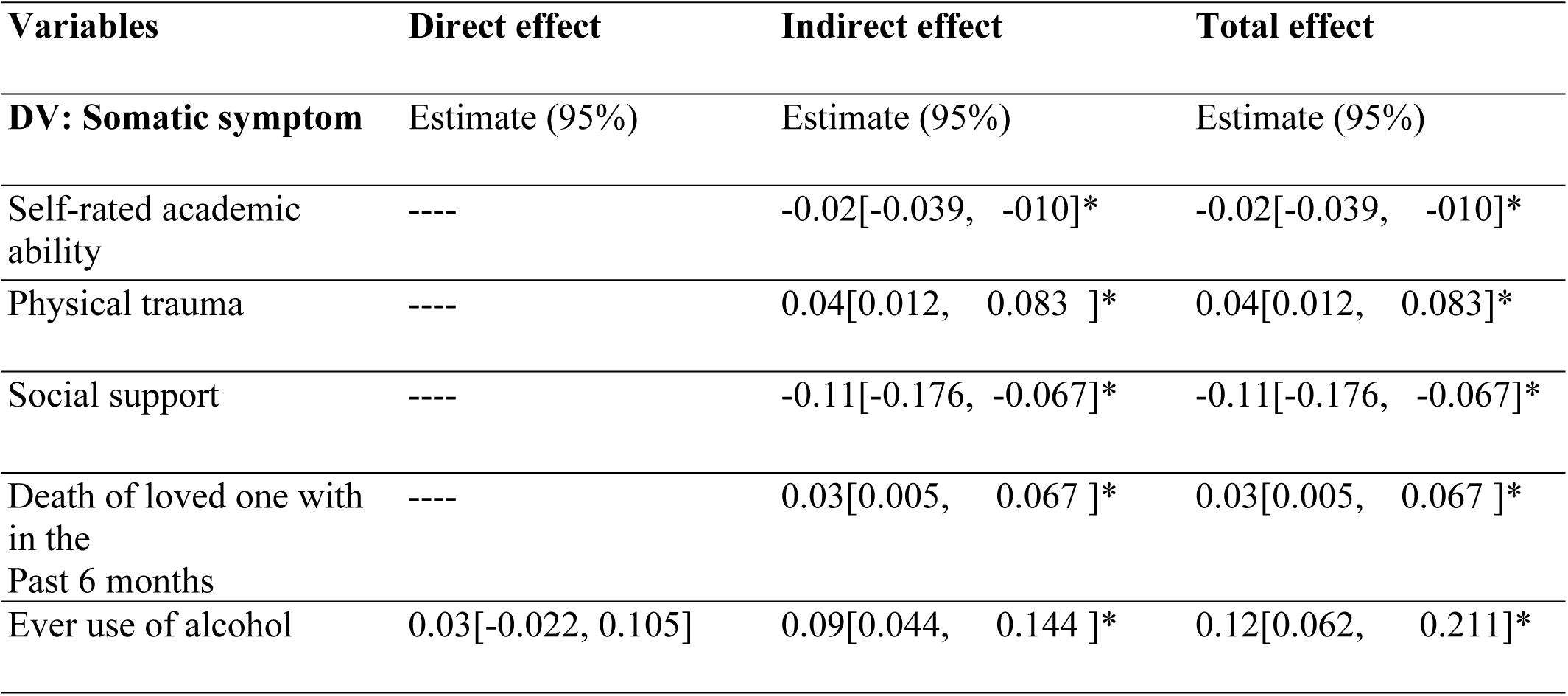
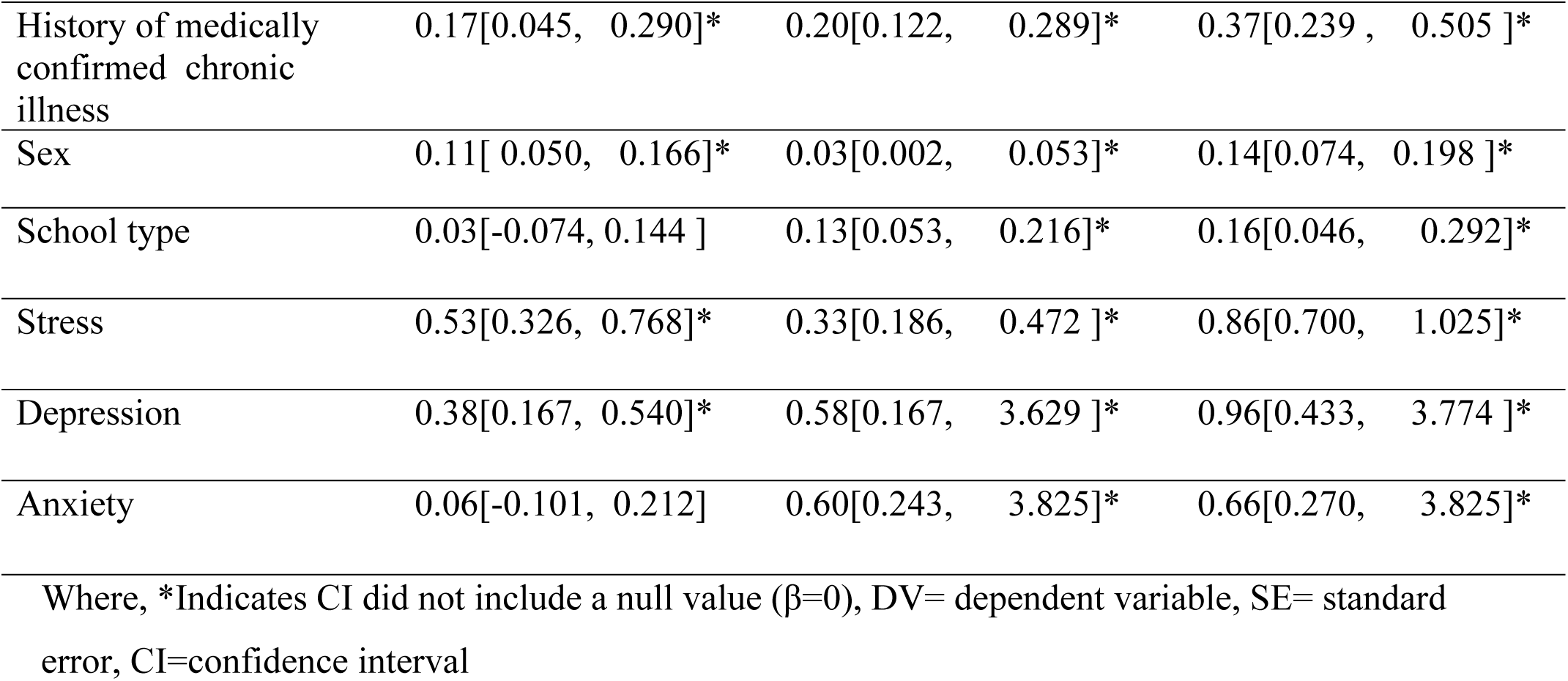
direct, indirect and total effects of socio-demographic, personal, relationship related factors, stress, anxiety, and depression on somatic symptom disorders among adolescents in Gondar town, Northwest Ethiopia, 2022. **Unstandardized estimate**.

### Factors related to anxiety among high and preparatory school adolescents

School type, sex, stress, and depression had statistically significant direct and indirect relationships with anxiety. Self-rated academic ability and physical trauma had direct effects, while ever use of alcohol, and having medically confirmed chronic illness had a statistically insignificant direct, but significant indirect effects on anxiety. Perceived social support, perceived family academic pressure, and death of beloved one within the past 6 months.

Self-rated academic ability had a direct negative effect [adjusted β= -0.03, 95% CI: -0.065, - 0.006] on anxiety. Having a history of physical trauma had positive effect [adjusted β= 0.06, 95% CI: 0.014, 0.125] on anxiety. Being female had a significant direct [adjusted β = 0.08, 95% CI: 0.019, 0.135] and indirect [adjusted β = 0.04, 95% CI: 0.004, 0.092] positive effect on anxiety with a total positive effect of 0.12 [adjusted β = 0.12, 95% CI: 0.059, 0.179]. Being in private school had direct [adjusted β= 0.12, 95% CI: 0.017, 0.226] and indirect [adjusted β= 0.16, 95% CI: 0.056, 0.269] positive effect on anxiety, leading to a total positive effect of 0.28 [adjusted β= 0.28, 95% CI: 0.159, 0.415]. Stress had both direct [adjusted β= 0.54, 95% CI: 0.293, 0.745] and indirect [adjusted β= 0.57, 95% CI: 0.379, 0.814] positive effect on anxiety, bringing a total positive effect of 1.10[adjusted β= 1.10, 95% CI: 0.955, 1.264]. Perceived social support had a significant negative indirect effect [adjusted β= -0.22, 95% CI: -.304, -0.139] on anxiety, while having a history of death of a loved one in the past 6 months had a significant positive indirect effect [adjusted β= 0.06, 95% CI: 0.010, 0.122]. Depression had positive direct effect [adjusted β= 0.74, 95% CI: 0.508, 1.010] on anxiety (*Table 8*).

### Factors related to depression among high and preparatory school adolescents in Gondar town, 2022

School type and anxiety were significantly related to depression both directly as well as indirectly. Perceived social support and having a history of death of a loved one within the past six months had significant direct effects on depression. While having medically confirmed chronic illness, having a history of alcohol use, perceived family academic pressure, sex, and stress had an insignificant direct, but significant indirect and total effects on depression. Self-rated academic ability, having history of physical trauma, and depression itself had a statistically significant indirect relationship with depression.

Perceived social support had a significant direct negative effect [adjusted β= -0.13, 95% CI: - 0.229,-0.029] on depression. History of experiencing death of beloved one within the past six months had a direct positive effect [adjusted β= 0.03, 95% CI: 0.014, 0.256] on depression. Learning private school had a significant negative direct [adjusted β = -0.09, 95% CI: -0.194, - 0.004] and positive indirect effect on depression [adjusted β = 0.2, 95% CI: 0.113, 0.320], with a total positive effect of 0.11 [adjusted β = 0.10, 95% CI: 0.016, 0.207]. Having a medically confirmed chronic illness had a significant indirect effect on depression [adjusted β = 0.17, 95% CI: 0.085, 0.265]. Self-rated academic ability had a statistically significant negative indirect effects [adjusted β=-0.05, 95% CI: -0.083,-0.031] on depression. Having a history of physical trauma had a statistically significant positive indirect effects [adjusted β= 0.10, 95% CI: 0.037, 0.166] on depression. Anxiety was significantly related to depression [adjusted β= 0.74, 95% CI: 0.483, 1.081] (***Table 9***).

### Factors related to somatic symptoms among high and preparatory school adolescents

Having a medically confirmed chronic illness, sex, stress, and depression were all directly and indirectly related to somatic symptoms. Having a history of alcohol drinking, school type and anxiety had insignificant direct, but significant indirect and total effects on somatic symptoms. Self-rated academic ability, perceived social support, physical trauma, and death of a loved one in the past 6 months had a significant indirect effects on somatic symptoms.

Having medically confirmed chronic illness had significant positive direct [adjusted β= 0.17, 95% CI= 0.045, 0.290] and indirect [adjusted β= 0.20, 95% CI: 0.122, 0.289] effect on somatic symptoms, bringing a total positive effects of 0.37 [adjusted β= 0.37, 95% CI: 0.239, 0.505]. Being female had statistically significant direct [adjusted β= 0.10, 95% CI: 0.050, 0.166] and indirect [adjusted β= 0.03, 95% CI: 0.002, 0.053] positive effect on somatic symptoms, resulting in a total positive effects of 0.13 [adjusted β= 0.13, 95% CI: 0.074, 0.198]. Stress had both direct [adjusted β= 0.53, 95% CI: 0.326, 0.768] and indirect [adjusted β= 0.33, 95% CI: 0.186, 0.472] positive effect on somatic symptoms resulting in a total positive effect of 0.86 [adjusted β= 086, 95% CI: 0.700, 1.025]. Depression had significant direct [adjusted β= 0.38, 95% CI= 0.167, 0.540] and indirect [adjusted β= 0.58, 95% CI: 0. 0.167, 3.629] positive effect on somatic symptoms resulting in a total positive effect of 0.96 [adjusted β= 0.96, 95% CI: 0.433, 1.456]. A history of alcohol consumption had a statistically insignificant direct effect on somatic symptoms [adjusted = 0.03, 95% CI: -0.022, 0.105], but it had a significant positive indirect effect [adjusted = 0.09, 95% CI: 0.044, 0.144], resulting in a total positive effect of 0.12 [adjusted = 0.12, 95% CI: 0.062, 0.211]. Anxiety has statistically insignificant direct effect on somatic symptoms [adjusted β= 0.06, 95% CI: -0.101, 0.212], however, it had statistically significant positive indirect effect [adjusted β= 0.6, 95% CI: 0.243, 3.825], resulting in a total positive effects of 0.66 [adjusted β=0.66, 95% CI: 0.270, 3.825]. Social support [adjusted = -0.11,% CI: -0.176, -0.067] and self-rated academic ability [adjusted =-0.02, 95%CI: -0.039, -010] had a significant negative indirect effect on somatic symptoms, whereas a history of death of a loved one within the previous six months [adjusted = 0.03% CI: 0.005, 0.067] and having history of physical trauma [adjusted β= 0.04, 95% CI:0.012, 0.083] had significant positive indirect effect on somatic symptom (***Table 10***).

## Discussion

In this study, the magnitude and determinants of anxiety, depression, and somatic symptoms were examined using non-recursive structural equation modeling. Magnitude of depression, anxiety, and somatic symptom were 28.21 [95% CI: 25.8, 31%], 35.82 [(95% CI: 33.3, 38.4%) and 25.24[95% CI: 23, 27.6%] respectively. Self-rated academic ability, physical trauma, school type, sex, stress, ever use of alcohol, perceived social support, death of a loved one, and having medically confirmed chronic illness were independent predictors of anxiety, depression and somatic symptoms. The bidirectional relationship between anxiety and depression was significant. Depression and anxiety were significant predictors of somatic symptom.

### Magnitude of depression and its associated factors

In the current study, the overall prevalence of self-reported depression was 28.21(95% CI= 25.8%, 31%). This is in line with the study conducted in Jimma 28% [18], but it was higher than meta-analysis conducted in china 24.3% [14], and lower than the study conducted in Aksum 38.2% [19], Nepal 44.2% [113], Bangladesh 36%[114], and India 57.7% [115]. The reason for this discrepancy may be due to differences in the screening tools used. Meta-analysis conducted in china included articles with different tools such as Beck Depression Inventory (BDI), Zung self-rating depression scale (SDS), and center of Epidemiology studies depression scale (CES-D). The study Nepal and India (CES-D). This variation could also be justified by the difference in the cutoff points used; the study conducted in Aksum and in Bangladesh used a cutoff above five. Furthermore, this could be explained by socio-cultural differences in different settings.

In the final model of our study, perceived social support, having a history of death of a loved one within the past six months, school type, sex, anxiety, having medically confirmed chronic illness, having a history of alcohol use, perceived family academic pressure, Self-rated academic ability and having a history of physical trauma had a statistically significant relationship with depression.

Being female had a statistically significant positive effects [adjusted β= 0.06, 95% CI:0.003, 0.109] on depression; this implies that being females increase the level of level of depression compared to males, controlling other factors constant. This finding is consistent with other studies [18, 37–39]. This could be explained by the fact that men and women have different brain chemistry and hormonal variations. Menstrual fluctuations in progesterone and estrogen may affect neurotransmitters in women, and low neurotransmitters may lead to depression[121]. Besides, adolescent girls are more likely to have negative life events in relation to their parents and peers, and females are more emotion focused and have a distracting coping style than males [122] . Furthermore, females are more susceptible to the opinions of others, which may leads to depression [123].

Perceived social support had a significant direct negative effect [adjusted β= -0.13, 95% CI: - 0.229,-0.029] on depression. This means that as social support increases, the level of depression decreased or students with low social support had higher levels of depression as compared to those who had high social support. This finding is compatible with different reports [43, 44, 52, 56]. The probable reason for this might be low social support which may increase feelings of loneliness, worthlessness, and hopelessness; leading to loss of interest in activities and depression. Moreover, students with low social support are more likely to have low self-esteem and negative cognition than those with higher social support, which may lead to depression.

Ever use of alcohol had a positive direct effect [β= 0.14, 95% CI: 0.073, 0.201] on depression. This implied that an adolescent who had a history of alcohol drinking had a higher level of depression as compared to who did not drink. This finding is congruent with the study conducted in USA [45] and in China [46]. This could be due to the fact that alcohol affects brain chemicals like serotonin and dopamine, which are responsible for happiness, and the decrease in thus chemicals leads to feeling down and depressed [124].

Students with lower self-rated academic ability had a higher levels of depression [adjusted β= -0.05, 95% CI= -0.083,-0.031] as compared with those with high self-rated academic ability. This result is compatible with the study conducted in Iran [125]. The possible explanation for this might be, students with low academic ability are terrified of negative responses from teachers, parents, and friends, which may lead to fear about their future career, they will lose confidence in managing their own problems, which will lead them to become anxious, and anxiety will lead to depression.

Stress had significant positive effect [adjusted β= 0.76, 95% CI: 0.642, 0.900] on depression. This indicates that participants with high-level stress had a higher levels of depression, holding other predictors constant. Our finding is supported with different literatures [49, 50]. The possible justification for this could be owing to the fact that high-level stress impairs the brain function, which may lead to fluctuations in neurotransmitters such as dopamine, which leads to depression. In addition, immune dysregulation during stress-full life events leads to anxiety and anxiety leads to depression.

Holding other factors constant, perceived family academic pressure had statistically significant positive effects [adjusted β= 0.04, 95% CI: 0.016, 0.060] on depression. Implies that participants with higher family academic pressure were related to higher level of depression. This finding is supported by a study conducted in Nepal [41] and in Vietnamese [42]. This could be the higher expectation of parents with higher academic pressure leading the student to stress which in turn leads to anxiety and anxiety leads to depression. Furthermore, students who fail to meet expectations may face harsh criticism, which may lead to self-doubt about their abilities, which may lead to a sense of worthlessness and hopelessness.

A history of experiencing death of beloved one within the past six months had a direct positive effect [adjusted β= 0.03, 95% CI: 0.014, 0.256] effect on depression, keeping other predictors constant. This suggests that students who had a history of death of a loved one in the past 6 months had higher levels of depression as compared with their counterparts. Possible justification for this could be individuals who have lost a loved one, may feel worthless without that person, and they may experience feelings of loneliness and poor social interaction, which can lead to depression[126]. Moreover, the emotional reaction following grave may lead to depletion of dopamine, which may lead to depression.

Participants with a history of physical trauma had a higher levels of depression [adjusted β= 0.10, 95% CI: 0.037, 0.166] compared to those who had not a history of trauma, holding other predictors constant. The plausible explanation for this could be individuals with physical trauma may have subsequent negatives teaches about the event, they may fear that the events will happen again, which may lead them to anxiety, and anxiety leads to depression.

Learning in a private school had statistically significant positive effects [adjusted β= 0.10, 95% CI: 0.016, 0.207] on depression. It can be inferred that participants in private schools had a higher levels of depression compared to their counter parts. In contrast to our finding, the study conducted in India [40] showed that public school students had a higher level of depression than private school students; this disparity may be explained by differences in sample size and method of analysis; our study had a larger sample size than the India study, and the India study did not address the confounding effect. Higher levels of depression in private schools may be due to parents’ expectations of high academic achievement in private schools; which lead students to be stressed when they try to meet their parents’ expectations, and stress leads to depression.

Holding other predictors constant, medically confirmed chronic illness increased the level of depression [adjusted β= 0.21, 95% CI: 0.114, 0.311]. This could be justified by the fact that chronic illness decreases the ability to do things, and people with chronic illness may fear the consequences of their illness. Which can lead to anxiety and anxiety can lead to depression. Furthermore, it can be explained by the physical effect of the condition and the treatment effect for those who are on treatment.

High level of anxiety was significantly related to higher level of depression [adjusted β= 0.74, 95% CI: 0.483, 1.081]. This finding is compatible with other findings [54, 55]. The possible justification for this may be an individual with anxiety may try to control their worries, but if they are unable to do so, their emotions will be affected and they will become down, hopeless, and depressed. It can also be explained by the sharing of a common set of genes and environmental factors [127].

### Magnitude of Anxiety and its associated factors

In this study, the prevalence of anxiety was 25.05% (95%CI: 22.8, 27.5%). It is consistent with the study conducted among children’s and youth in Ethiopia 0.5 to23% [28]. However, this finding is lower than the study conducted in Kenya 37.99% [17], Saudi Arabia 63.5% to 66% [116, 117], in Jordan 42.1% [26], and in Chandigarh 80.85% [27]. In contrast to this, it was higher than the worldwide estimates of anxiety among children and adolescents (6.5 to 10 %) [118], and among the study conducted in 82 countries, which reported 13-14% in Africa, 9-10% in America, 8-9% in Europe, and 7-8% in Asia [25]. Such variation in the prevalence of anxiety across different continents could originate from variations in the anxiety screening tools used. A study conducted in Saudi Arabia used depression anxiety stress scale (DASS-42, a study conducted in Jordan used symptom checklists anxiety scale, a study conducted in Chandigarh used depression anxiety stress scale-21. Besides, it could also be justified by the different in the cutoff point used: a study conducted in Saudi Arabia used GAS-above five. Thus, variation may also be explained by deference in population characteristics, a study conducted in 82 countries community based study which may decrease the magnitude. Furthermore, cultural differences in different parts of the world may lead to such variations.

From the final non-recursive structural equation model our study, we found that self-rated academic ability and perceived social support had a significant negative effects on anxiety, depression, stress, physical trauma, school type, sex, ever use of alcohol, perceived family academic pressure, medically confirmed chronic illness, death of a loved one within the past 6 months had significant positive effect on anxiety.

As self-rated academic ability of the student increases, the level of anxiety decreases [adjusted β= -0.03, 95% CI: -0.065, -0.006]. This finding was supported by different studies [26, 59, 128]. The plausible reason for this might be students with high self-rated academic ability will be more likely to make decisions, express their feelings without feeling embarrassed, and be more optimistic. Moreover, it could be explained by encouragement and positive response from parents, teachers, and peers.

In a study conducted in china [129], alcohol has positive effect on anxiety. Which is supported by our study [adjusted β= 0.11, 95% CI: 0.041, 0.188]. It can be inferred that adolescents who had a history of alcohol drinking had a higher levels of anxiety than their counterparts, keeping other predictors constant. This could be owing to the fact that ethanol disrupts brain development by killing neurons, resulting in cognitive and affective dysfunction [130] .Which can lead to stress and depression, and stress and depression can lead to anxiety. Furthermore, alcohol drinking may lead to anxiety by decreasing the age-appropriate activities of adolescents such as physical activity and social relationships. Besides, alcohol increases conflict with others [131] and conflict leads to stress that may lead to anxiety.

Being female had a statistically significant positive effect [adjusted β= 0.12, 95% CI: 0.059, 0.179] on anxiety. This suggests that controlling other predictors’ constant, female students had higher levels of anxiety than male students. This finding is in harmony with the study conducted in different continents [27, 57]. The possible explanation could be that females may be subjected to more life stressors, such as the monthly menstrual cycle and psychological and physical violence [132]. In addition, it can be explained by the higher hormonal fluctuation in women, which exposes them to become anxious [133].

In the current study, perceived social support had a significant negative indirect effect [adjusted β=-0.22, 95% CI: -.304, -0.139] on anxiety. This implied that, controlling other determinants constant, as social support increases, the anxiety level will decrease. Direct Comparison is difficult, since none of the other studies assessed the indirect effect of social support on anxiety through depression. This may be explained by the fact that low social support increases the feeling of loneness and worthless. Which leads to hopelessness, low self-esteem, and negative cognition; that may lead to depression and depression leads to anxiety.

Stress had significant positive effects [adjusted β= 1.10, 95% CI: 0.955, 1.264] on anxiety. It can be concluded that, as the level of the level of stress increases, the level of anxiety will also increase, keeping other factors constant. Our finding is compatible with different literatures[60, 61]. This could be justified by the fact that high levels of stress cause immune dysregulation, which causes an increase in cortisol and corticotrophin hormones, which can impair brain function and eventually lead to anxiety.

Perceived family academic pressure had an indirect positive effect [adjusted β= 0.03, 95% CI: 0.001, 0.50] on anxiety. Means that, as student perceived family academic pressure increases, the level of anxiety will also increase. This finding is supported with the finding in Nepal [41], in south Africa [134], and in Vietnamese [42]. This might be justified by the fact that parents with high academic pressure have higher academic expectations, when students trying to cope with this high expectation may develop stress which in turns leads to anxiety [44].

Controlling other factors, constant death of a beloved one within the past 6 months had a significant positive indirect effect [adjusted β= 0.06, 95% CI: 0.010, 0.122] on anxiety. Means those participants who had a history of death of the beloved one in the past 6 months had a higher level of anxiety compared to their counterparts. This could be an individual with death of a loved one, may consider themselves as worthless without that person, they may develop a feeling of loneness, which might lead to depression and depression leads to anxiety.

Our findings showed that, holding other predictors constants, having a history of physical trauma had a direct positive effect [adjusted β= 0.06, 95% CI: 0.014, 0.125] on anxiety. This indicates that students with physical trauma had higher levels of anxiety as compared with those who had not. Our finding was a supported by the study conducted in Netherland [135]. This consistency could be justified by the fact that in reaction to trauma, neurotransmitters such as dopamine, serotonin, and gamma amino butyric acid may be impacted by cortisol, and depletion of thus neurotransmitters may lead to anxiety. Besides, individuals with physical trauma may internalize the event and may have subsequent negative thoughts about themselves; which affect their emotional experience, which might leads to anxiety [136].

A study conducted in India [137] reported that school type had not a significant relationship with anxiety. Contrarily, in our study school type had a significant relationship with level of anxiety [adjusted β= 0.28, 95% CI: 0.159, 0.415. It implied that adolescents in private schools experienced higher levels of anxiety than their peers did. It could be due to high control and supervision by parents and teachers in private schools. Furthermore, it might be due to the high academic expectation of students in private schools to satisfy their parents, which may lead to become anxious by fearing future fear of low academic achievement.

Having medically confirmed chronic illness had significant positive effects [adjusted β= 0.25, 95% CI: 0.132, 0.371] on anxiety. The plausible explanation for this could be that individuals with chronic illness may have excessive fear about it consequences; they may worry about what peoples say about their illness. This leads to anxiety. Furthermore, it could be justified by the physical effect of the condition, the treatment effect for those who started treatment, which may directly lead to depression, and depression leads to anxiety.

Controlling other predictors, constant depression had a direct positive effect on anxiety [adjusted β= 0.74, 95% CI: 0.508, 1.010]. Our finding is in an agreement with the study conducted in United Kingdom [63]. This could be depressed people spend a lot of time worrying about their symptoms, which causes anxiety. Moreover, it could be due to a shared set of genes [127].

### Somatic symptom and its associated factors

In the current analysis, the prevalence of self-reported somatic symptom disorder was 25.24 % (95% CI= 23, 27.6%). This is consistent with the study conducted among children and adolescents in Tarragona which reported 5 to 30 % [35]. The finding in the present study was higher than the magnitude in the general population 5 to 7% [119] and lower than study conducted in Qatar, which reported 47.8% in female and 52.2% in male [120]. The possible explanation for this could be due to variation in the screening tools. The study conducted in Qatar used General Health Questionnaire-12. The study among the general population of children and adolescents used a child somatization inventory. Furthermore, it could also be a variation in population, the study conducted in Qatar was conducted among patients and the study among the general population of children and adolescents including non-school adolescents. Moreover, socio-cultural variation could also be the possible justification.

#### Factors related with depression, anxiety, and somatic symptoms

In the analysis results of the current study, medically confirmed chronic illness, sex, stress, and depression were significantly related with somatic symptoms both directly and indirectly. Having a history of alcohol drinking, school type, anxiety, self-rated academic ability, perceived social support, physical trauma, and death of the beloved one within the past 6 months had a significant indirect effects on somatic symptoms.

Controlling other predictors, constants, medically confirmed chronic illness had a significant positive effect [adjusted β= 0.37, 95% CI: 0.239, 0.505 on somatic symptoms. This could be individuals with chronic illness may have excessive fear about it consequence which leads to anxiety and anxiety leads to a somatic symptoms. Moreover, it could be by the physical effect of the condition and treatment effect for those who are on treatment, which may directly lead to depression, and depression leads to a somatic symptoms. Direct effect of chronic illness on cognitive function may also be the possible reason.

Keeping other factors, constant the history of death of the beloved one in the past 6 months had a significant indirectly effects [adjusted β= 0.03,% CI: 0.005, 0.067] on somatic symptoms. Direct comparison is not possible, since there is no study conducted with adolescents in Ethiopia or outside. But, it is consistent with the study conducted in Nepal among widows [138]. The possible explanation could be an individual with death of beloved, one may consider themselves as worthless without that person, they could have a feeling of loneness and poor social interaction which leads to depression [126] and depression leads to somatic symptoms.

Being female had a statistically significant positive effect [adjusted β=0.13, 95% CI: 0.074, 0.198] on somatic symptoms. This suggests that female students had higher levels of somatic symptoms compared to male students, holding other determinants constant. This finding is supported with other studies [64, 65]. The possible justification for consistency could be psychosocial and biological factors or complex mixture of them [122] that may lead women’s at risk of somatic symptoms. Moreover, it could be justified by women’s ability to quickly acknowledge psychological distress and seek help for it.

As the stress level increase, the level of somatic symptoms will also increase [adjusted β= 086, 95% C: 0.700, 1.025]. This means that students with lower levels of stress had a lower levels of somatic symptoms compared to those with lower levels of stress. Our finding is in line with the study conducted in Sweden [70]. The plausible explanation for this might be high cortisol release in response to stress may affect neuro-transmitters; which could lead to impairment in brain function and development of somatic symptom disorders.

As the level of depression increase, the level of somatic symptoms will also increase [adjusted β= 0.96, 95% CI: 0.167, 3.629], which means that students with higher levels of depression had a higher levels of somatic symptoms. This result is supported with other studies [65, 75, 76]. Possible explanation for this consistency could be in the depressed individual’s neurotransmitters like nor-epinephrine and serotonin are low and the decrement in such neurotransmitters may lead individuals to focus on physical symptoms. Moreover, it could be explained by the co-occurrence of somatic symptoms and depression. Sharing of common sets of genes and environmental factors may also be a possible justification.

In our study, having ever use of alcohol had a statistically significant positive effect [adjusted β= 0.12, 95% CI: 0.062, 0.211] on somatic symptoms, keeping other predictors constant; this means that individuals who had a history of alcohol use had a higher levels of somatic symptom compared to their counterparts. Compatible with our finding, a literature review on the relationship between anxiety, depression and substance use [139] showed similar finding. The probable reason could be the direct effect of ethanol on nerve cell [130] which leads to heightened awareness of normal body sensation.

Anxiety had significant positive effect [adjusted β=0.66, 95% CI: 0.270, 3.825] on somatic symptoms. This implies that participants with a higher level of anxiety will have higher level of somatic symptom as compared to those with lower levels of anxiety. The result of the present study is line with other studies [65, 73, 74]. It could be excessive and uncontrolled, worrying in an anxious individual may lead to depression, and depression in turn leads to a somatic symptoms. Besides, a common set of gene components for anxiety and somatic symptoms might also be the possible reason.

Social support had a statistically significant indirect negative effect [adjusted β=-0.11,% CI: -0.176, -0.067] on somatic symptoms; implying that as social support increases the level of somatic symptoms will also increase. In other words, students with low social support had a higher level of somatic symptoms, compared to those with high social support. This finding is congruent with the study conducted in United Kingdom [140]. It could be justified by individuals with low social support may have low self-esteem and negative cognition, which could lead to depression and depression could lead to somatic symptoms.

Self-rated academic ability had a significant negative indirect effect [adjusted β= -0.02, 95%% CI: -0.039, -010] on somatic symptoms; implying that holding other predictors constant, as student self-rated academic ability increased the level of somatic symptoms will also increase. The present study was supported by the study conducted in Slovakia [138].The possible explanation for this consistency could be students spending half of their day time in school. For students with lower academic achievement, interaction with teachers and with their peers will be affected; which leads to anxiety and anxiety leads to a somatic symptoms.

Having a history of physical trauma had a significant positive indirect effect [adjusted β= 0. 04, 95% CI: 0.012, 0.083] on somatic symptoms. It can be interpreted as school adolescents with a history of physical trauma had a higher level of somatic symptoms as compared to their counterparts. This finding is consistent with other study [141]. This may be justified by the fact that trauma could have long-lasting effects on how individuals recognize themselves and their coping abilities, which leads to anxiety and in turn anxiety leads to somatic symptoms. Moreover, new stressful events may intensify the effect of trauma, which will aggravate already present somatic symptoms.

Being in a private schools increased the level of somatic symptoms [adjusted β= 0.16, 95% CI: 0.046, 0.292] compared to being in public schools, controlling other factors constant. This finding is consistent with the study conducted in Turkey[64]. Possible justification could be private school students and their parents are optimistic of academic achievement than public school students and parents. Great expectation of high academic achievement leads to the creation of high standards of behavior, which may lead them to become anxious and anxiety leads to a somatic symptoms. Furthermore, this may be due to higher parental and teacher supervision and control in private schools which lead to low psychological autonomy.

## Strength and limitations

Bidirectional relationship between anxiety and depression was assessed. Besides, the indirect and direct effects of different independent variables on different dependent variables were assed simultaneously. However, this study is not without limitations, only school adolescents were included in the study, which may affect the generalizability to all adolescents.

## Conclusion

In this study, a moderate magnitude of anxiety, depression, and somatic symptoms were found. Stress, school type, and sex had significant direct and indirect relationship with anxiety.

Depression, self-rated academic ability and physical trauma had a direct effects, while ever use of alcohol, medically confirmed chronic illness, perceived social support, perceived family academic pressure, and death of a loved one within the past 6 months had a significant indirect relationship with anxiety.

School type and anxiety were significantly related with depression both directly and indirectly. Perceived social support and having a history of death of a loved one within the past six months had statistically significant direct effect on depression. Whereas, having medically confirmed chronic illness, ever use of alcohol, perceived family academic pressure, sex, stress, self-rated academic ability, and having a history of physical trauma had a significant indirect relationship with depression.

Depression, stress, medically confirmed chronic illness, and being female were significantly related with somatic symptoms both directly and indirectly. Simultaneously, alcohol use in the past 6 months, school type, anxiety, self-rated academic ability, perceived social support, physical trauma, and death of a loved one had a significant indirect effect on somatic symptoms.

## Data Availability

All relevant data are within the manuscript and its Supporting Information files

## Abbreviations

AIC: Akaike information Criteria
OVB: Omitted variable bias
CFI: Comparative Fix index
CMB: Common Method Bias
GBD: Global Burden of Disease
GFI: Goodness of Fit Index
IRB: Institutional Review Board
IV: Instrumental Variable
MD: Mahalanobis distance
OVB: Omitted Variable Bias
PHQ-9A: Patent Health Questionnaire for Adolescent
PSS: Perceived Stress Scale
RMSEA: Rooted Mean Square Error of Approximation
SDG: Sustainable Development Goal
SSS: Somatic Symptom Scale
TLS: Tucker Lewis Index
WHO: World Health Organization

## Acknowledgments

First and foremost, we would like to express our gratitude to the Almighty God, who has served as our compass and source of strength throughout the paper. Next, we would like to thank University of Gondar College of Medicine and Health Science, Institute of Public Health for giving us this great opportunity to do this paper and for the internet access. Following that, we would like to thank the Gondar Town Administrative Education Department and the School Directors for their information. Finally, we would like to thank each participant for agreeing to take part and providing the essential data.

## Authors’ contribution

Conceptualization: Zenebe Abebe Gebreegziabher

Data curation: Zenebe Abebe Gebreegziabher. Ayenew Molla, Rediet Eristu Formal analysis: Zenebe Abebe Gebreegziabher

Investigation: Zenebe Abebe Gebreegziabher. Ayenew Molla, Rediet Eristu Methodology: Zenebe Abebe Gebreegziabher. Ayenew Molla, Rediet Eristu

Project administration: Zenebe Abebe Gebreegziabher. Ayenew Molla, Rediet Eristu Resource: Zenebe Abebe Gebreegziabher. Ayenew Molla, Rediet Eristu

Software: Zenebe Abebe Gebreegziabher, Ayenew Molla, Rediet Eristu Supervision: Zenebe Abebe Gebreegziabher, Ayenew Molla, Rediet Eristu Validation: Zenebe Abebe Gebreegziabher, Ayenew Molla, Rediet Eristu Visualization: Zenebe Abebe Gebreegziabher

Writing – original draft: Zenebe Abebe Gebreegziabher

Writing – review and editing: Zenebe Abebe Gebreegziabher. Ayenew Molla, Rediet Eristu

## Supporting Information’s

S1 Table1 1: tools

S1 Table1 2: intraclass correlation coefficients

S1 Table1 3: Instrumental variable validity and strength

S1 Table1 4: depression among adolescents

S1 Table1 5: anxiety among adolescents

S1 Table1 6: somatic symptom among adolescent S1 Fig 1: hypothetical measurement model

S1 Table1 7: Factorial average item parceling S2 1: Data set

## Notes

### Competing Interest Statement

The authors have declared no competing interest.

### Funding Statement

The author(s) received no specific funding for this work.

### Author Declarations

Ethical approval was obtained from the Institutional Review Board (IRB) of University of Gondar Institute of public Health. The ethical approval letter was submitted to Gondar town administrative educational department and to all selected high schools and preparatory schools. Permission was obtained from school directors. Following permission from school directors, a detailed participant information sheets were given to each student, and they completed an assent form to indicate a willingness to participate.

